# Systematic Review of Large Language Models for Patient Care: Current Applications and Challenges

**DOI:** 10.1101/2024.03.04.24303733

**Authors:** Felix Busch, Lena Hoffmann, Christopher Rueger, Elon HC van Dijk, Rawen Kader, Esteban Ortiz-Prado, Marcus R Makowski, Luca Saba, Martin Hadamitzky, Jakob Nikolas Kather, Daniel Truhn, Renato Cuocolo, Lisa C Adams, Keno K Bressem

## Abstract

The introduction of large language models (LLMs) into clinical practice promises to improve patient education and empowerment, thereby personalizing medical care and broadening access to medical knowledge. Despite the popularity of LLMs, there is a significant gap in systematized information on their use in patient care. Therefore, this systematic review aims to synthesize current applications and limitations of LLMs in patient care using a data-driven convergent synthesis approach. We searched 5 databases for qualitative, quantitative, and mixed methods articles on LLMs in patient care published between 2022 and 2023. From 4,349 initial records, 89 studies across 29 medical specialties were included, primarily examining models based on the GPT-3.5 (53.2%, n=66 of 124 different LLMs examined per study) and GPT-4 (26.6%, n=33/124) architectures in medical question answering, followed by patient information generation, including medical text summarization or translation, and clinical documentation. Our analysis delineates two primary domains of LLM limitations: design and output. Design limitations included 6 second-order and 12 third-order codes, such as lack of medical domain optimization, data transparency, and accessibility issues, while output limitations included 9 second-order and 32 third-order codes, for example, non-reproducibility, non-comprehensiveness, incorrectness, unsafety, and bias. In conclusion, this study is the first review to systematically map LLM applications and limitations in patient care, providing a foundational framework and taxonomy for their implementation and evaluation in healthcare settings.

## 1. Introduction

Public and academic interest in large language models (LLMs) and their potential applications has increased substantially, especially since the release of OpenAI’s ChatGPT (Chat Generative Pre-trained Transformers) in November 2022.^1–3^ One of the main reasons for their popularity is the remarkable ability to mimic human writing, a result of extensive training on massive amounts of text and reinforcement learning from human feedback.^4^

Since most LLMs are designed as general-purpose chatbots, recent research has focused on developing specialized models for the medical domain, such as Meditron or BioMistral, by enriching the training data of LLMs with medical knowledge.^5,6^ However, this approach to fine-tuning LLMs requires significant computational resources that are not available to everyone and is also not applicable to closed-source LLMs, which are often the most powerful. Therefore, another approach to improve LLMs for biomedicine is to use techniques such as Retrieval-Augmented Generation (RAG).^7^ RAG allows information to be dynamically retrieved from medical databases during the model generation process, enriching the output with medical knowledge without the need to train the model.

LLMs hold great promise for improving the efficiency and accuracy of healthcare delivery, e.g., by extracting clinical information from electronic health records, summarizing, structuring, or explaining medical texts, streamlining administrative tasks in clinical practice, and enhancing medical research, quality control, and education.^8–10^ In addition, LLMs have been shown to be versatile tools for supporting diagnosis or serving as prognostic models.^11,12^

In contrast to applications primarily aimed at healthcare professionals, LLMs could also be used to promote patient education and empowerment by providing answers to medical questions and translating complex medical information into more accessible language.^4,13^ Thereby, LLMs may promote personalized medicine and broaden access to medical knowledge, empowering patients to actively participate in their healthcare decisions.

However, despite the growing body of research and the clear potential of LLMs, there is a gap in terms of systematized information towards their use in patient care. To date, there has been no evaluation of existing research to understand the scope of applications and identify limitations that may currently limit the successful integration of LLMs into clinical practice.

Therefore, this systematic review aims to analyze and synthesize the literature on LLMs in patient care, providing a systematic overview of 1) current applications and 2) challenges and limitations, with the purpose of establishing a foundational framework and taxonomy for the implementation and evaluation of LLMs in healthcare settings.

## 2. Methods

This systematic review was pre-registered in the International Prospective Register of Systematic Reviews (PROSPERO) under the identifier CRD42024504542 before the start of the initial screening and was conducted according to the Preferred Reporting Items for Systematic Reviews and Meta-Analyses (PRISMA) guidelines.^14,15^

### 2.1 Eligibility criteria

We searched 5 databases, including the Web of Science, PubMed, Embase/Embase Classic, American for Computing Machinery (ACM) Digital Library, and Institute of Electrical and Electronics Engineers (IEEE) Xplore as of January 25, 2024, to identify qualitative, quantitative, and mixed methods studies published between January 1, 2022, and December 31, 2023, that examined the use of LLMs for patient care. LLMs for patient care were defined as any artificial neural network that follows a transformer architecture and can be used to generate and translate text and other content or perform other natural language processing tasks for the purpose of disease management and support (i.e., prevention, preclinical management, diagnosis, treatment, or prognosis) that could be directly directed to or used by patients. Articles had to be available in English and contain sufficient data for thematic synthesis (e.g., conference abstracts that did not provide sufficient information on study results were excluded). Given the recent surge in publications on LLMs such as ChatGPT, we allowed for the inclusion of preprints if no corresponding peer-reviewed article was available. Duplicate reports of the same study, non-human studies, and articles limited to technology development/performance evaluation, pharmacy, human genetics, epidemiology, psychology, psychosocial support, or behavioral assessment were excluded.

### 2.2 Screening and data extraction

Initially, we conducted a preliminary search on PubMed and Google Scholar to define relevant search terms. The final search strategy included terms for LLMs, generative AI, and their applications in medicine, health services, clinical practices, medical treatments, and patient care (as detailed by database in Supplementary Section 1). After importing the bibliographic data into Rayyan and removing duplicates, LH and CR conducted an independent blind review of each article’s title and abstract.^16^ Any article flagged as potentially eligible by either reviewer proceeded to the full-text evaluation stage. For this stage, LH and CR used a custom data extraction form created in Google Forms (available online)^17^ to collect all relevant data independently from the studies that met the inclusion criteria. Quality assessment was also performed independently for each article within this data extraction form, using the Mixed Methods Appraisal Tool (MMAT) 2018.^18^ Disagreements at any stage of the review were resolved through discussion with the author FB. In cases of studies with incomplete data, we have tried to contact the corresponding authors for clarification or additional information.

### 2.3 Data analysis

Due to the diversity of investigated outcomes and study designs we sought to include, including qualitative, quantitative, and mixed methods, a meta-analysis was not practical. Instead, a data-driven convergent synthesis approach was selected for thematic syntheses of LLM applications and limitations in patient care.^19^ Following Thomas and Harden, FB coded each study’s numerical and textual data in Dedoose using free line-by-line coding.^20,21^ Initial codes were then systematically categorized into descriptive and subsequently into analytic themes, incorporating new codes for emerging concepts within a hierarchical tree structure. Upon completion of the codebook, FB and LH reviewed each study to ensure consistent application of codes. Discrepancies were resolved through discussion with the author KKB, and the final codebook and analytical themes were discussed and refined in consultation with all contributing authors.

## 3. Results

### 3.1 Screening results

Of the 4,349 reports identified, 2,991 underwent initial screening, and 126 were deemed suitable for potential inclusion and underwent full-text screening. Two articles could not be retrieved because the authors or the corresponding title and abstract could not be identified online. Following full-text screening, 35 articles were excluded, and 89 articles were included in the final review. Most studies were excluded because they targeted the wrong discipline (n=10/35, 28.6%) or population (n=7/35, 20%) or were not original research (n=8/35, 22.9%) (see Supplementary Section 2). For example, we evaluated a study that focused on classifying physician notes to identify patients without active bleeding who were appropriate candidates for thromboembolism prophylaxis.^22^ Although the classification tasks may lead to patient treatment, the primary outcome was informing clinicians rather than directly forwarding this information to patients. We also reviewed a study assessing the accuracy and completeness of several LLMs when answering Methotrexate-related questions.^23^ This study was excluded because it focused solely on the pharmacological treatment of rheumatic disease. For a detailed breakdown of the inclusion and exclusion process at each stage, please refer to the PRISMA flowchart in Figure 1.

**Figure 1.**
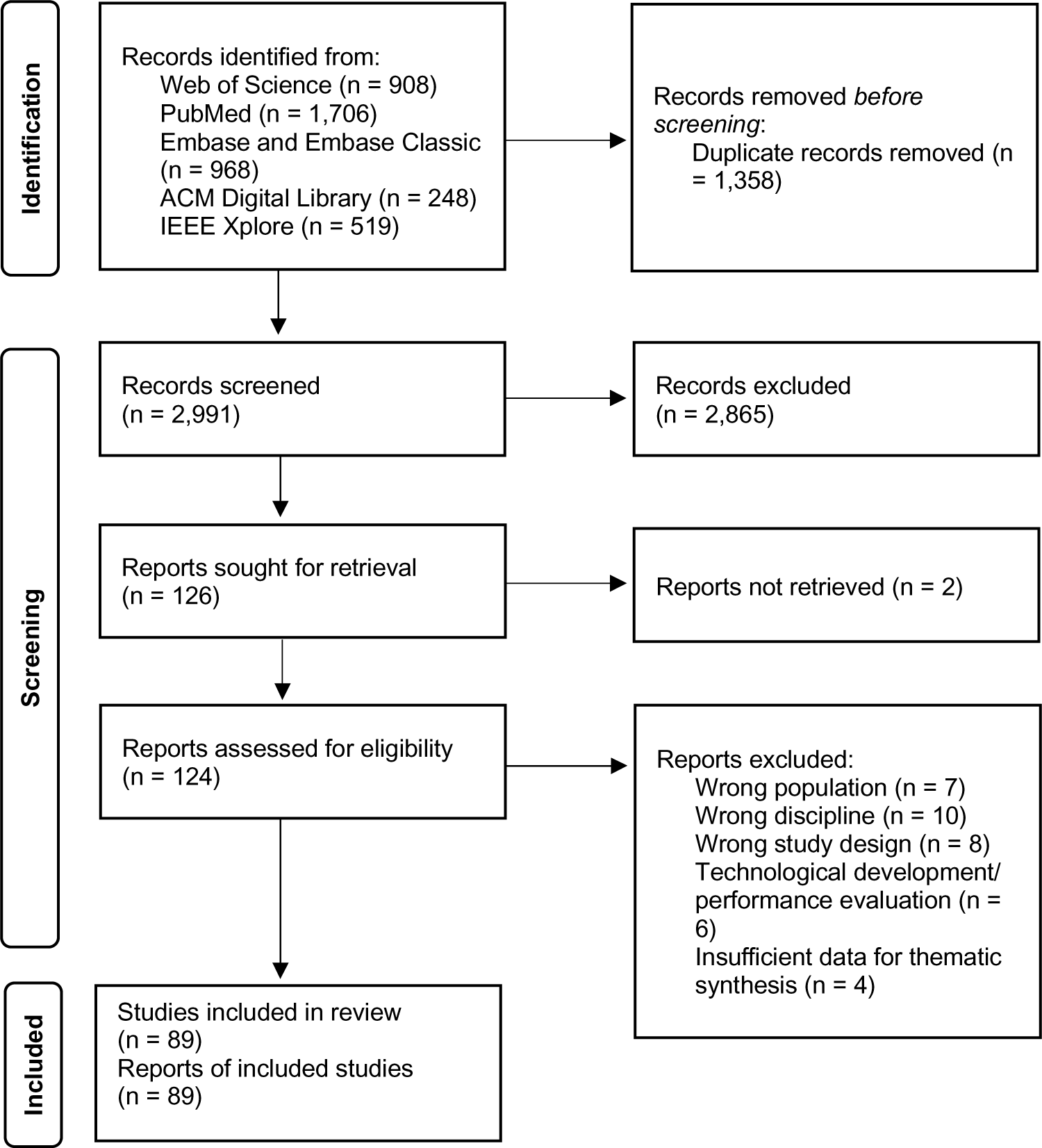
Preferred Reporting Items for Systematic Reviews and Meta-Analyses (PRISMA) flow diagram.

### 3.2 Characteristics of included studies

Table 1 summarizes the characteristics of the analyzed studies, including their setting, results, and conclusions. One study (n=1/89, 1.1%) was published in 2022^24^, 84 (n=84/89, 94.4%) in 2023^13,25–107^, and 4 (n=4/89, 4.5%) in 2024^108–111^ (all of which were peer-reviewed publications of preprints published in 2023). Most studies were quantitative non-randomized (n=84/89, 94.4%)^13,25–27,29–101,103,104,106,107,109–111^, 4 (n=4/89, 4.5%)^28,102,105,108^ had a qualitative study design, and one (n=1/89, 1.1%)^24^ was quantitative randomized according to the MMAT 2018 criteria. However, the LLM outputs were often first analyzed quantitatively but followed by a qualitative analysis of certain responses. Therefore, if the primary outcome was quantitative, we considered the study design to be quantitative rather than mixed methods, resulting in the inclusion of zero mixed methods studies. The quality of the included studies was mixed (see Table 2). The authors were primarily affiliated with institutions in the United States (n=47 of 122 different countries identified per publication, 38.5%), followed by Germany (n=11/122, 9%), Turkey (n=7/122, 5.7%), the United Kingdom (n=6/122, 4.9%), China/Australia/Italy (n=5/122, 4.1%, respectively), and 24 (n=36/122, 29.5%) other countries. Most studies examined one or more applications based on the GPT-3.5 architecture (n=66 of 124 different LLMs examined per study, 53.2%)^13,26–29,31–34,36–40,42–49,52–54,56–61,63,65–67,71,72,74,75,77,78,81–89,91,92,94,95,97–100,102–104,106–109,111^, followed by GPT-4 (n=33/124, 26.6%)^13,25,27,29,30,34–36,41,43,50,51,54,55,58,61,64,68–70,74,76,79–81,83,87,89,90,93,96,98,99,101,105^, Bard (n=10/124, 8.1%; now known as Gemini)^33,48,49,55,73,74,80,87,94,99^, Bing Chat (n=7/124, 5.7%; now Microsoft Copilot)^49,51,55,73,94,99,110^, and other applications based on Bidirectional Encoder Representations from Transformers (BERT; n=4/124, 3.2%)^13,83,84^, Large Language Model Meta-AI (LLaMA; n=3/124, 2.4%)^55^, or Claude by Anthropic (n=1/124, 0.8%)^55^. The majority of applications were primarily targeted at patients (n=64 of 89 included studies, 73%)^24,25,29,32,34–39,41–43,45–48,52–54,56–60,62,63,65,66,68–71,73–75,77–80,85–95,97,99,100,102–111^ or both patients and caregivers (n=25/89, 27%)^13,26–28,30,31,33,40,44,49–51,55,61,64,67,72,76,81–84,96,98,101^. Information about conflicts of interest and funding was not explicitly stated in 23 (n=23/89, 25.8%) studies, while 48 (n=48/89, 53.9%) reported that there were no conflicts of interest or funding. A total of 18 (n=18/89, 20.2%) studies reported the presence of conflicts of interest and funding.^13,24,38,40,54,58,59,67,69–71,74,80,84,96,103,105,111^ Most studies did not report information about the institutional review board (IRB) approval (n=55/89, 61.8%) or deemed IRB approval unnecessary (n=28/89, 31.5%). Six studies obtained IRB approval (n=6/89, 6.7%).^52,82,84–86,92^

**Table 1.**
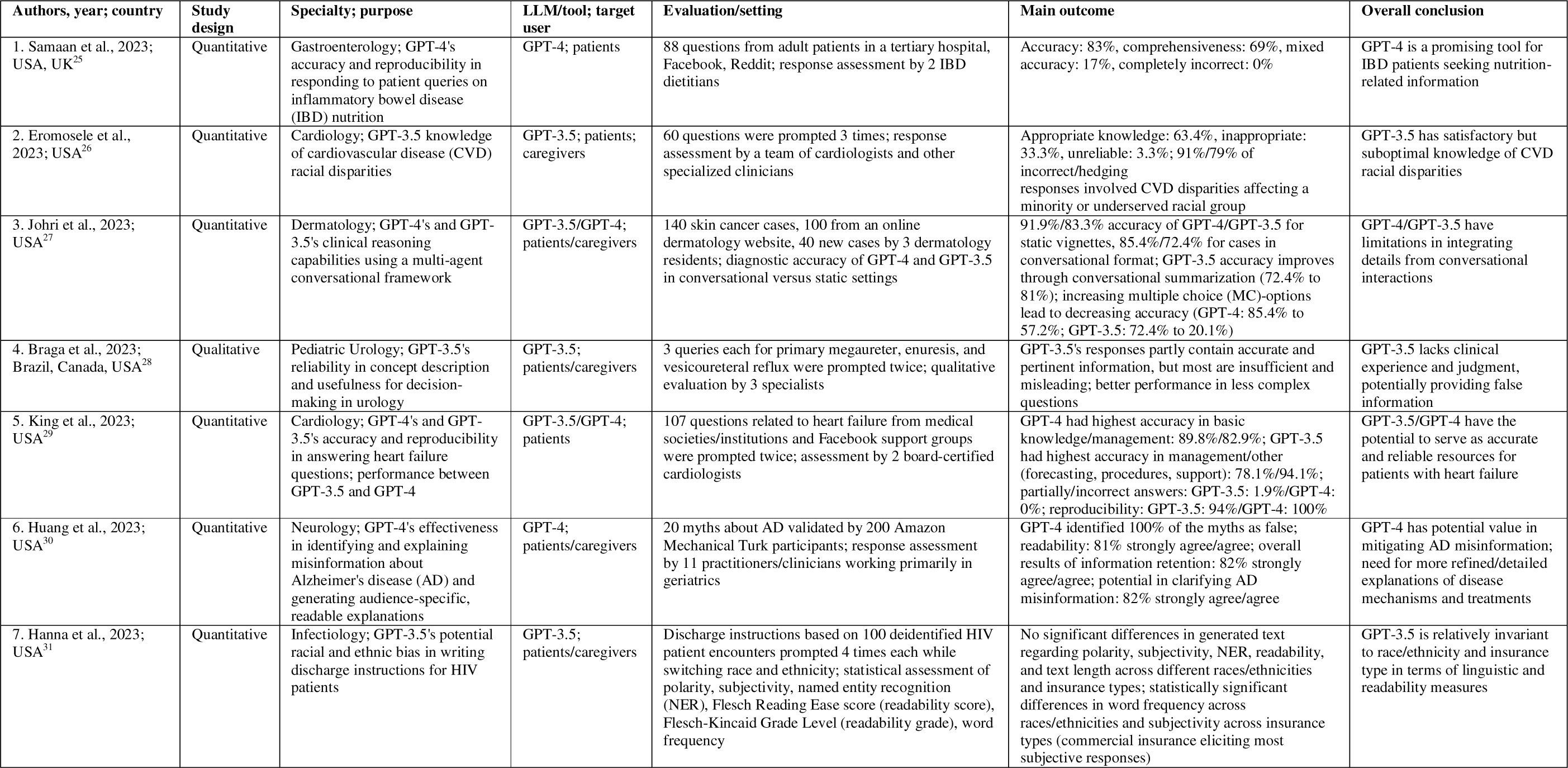
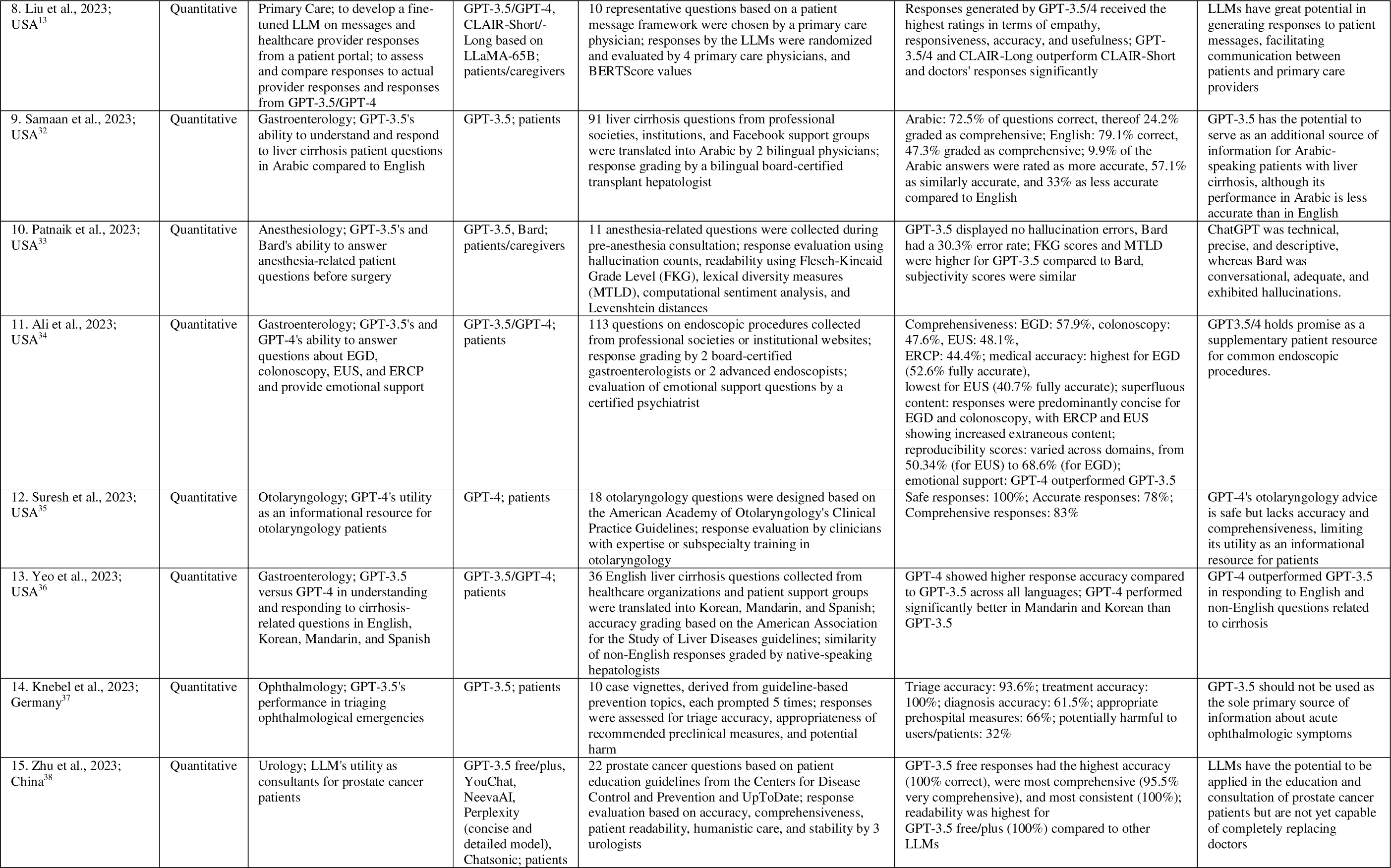
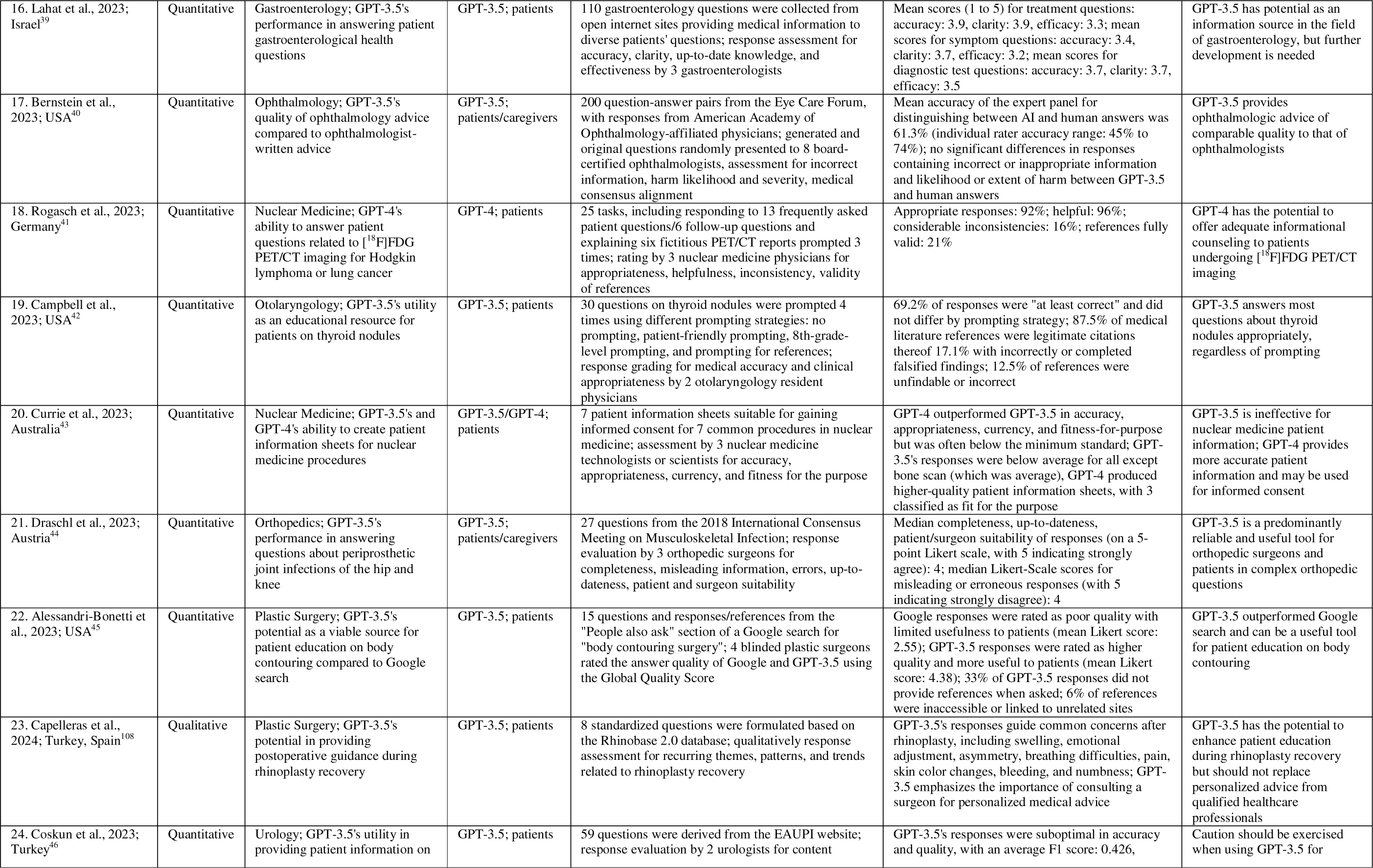
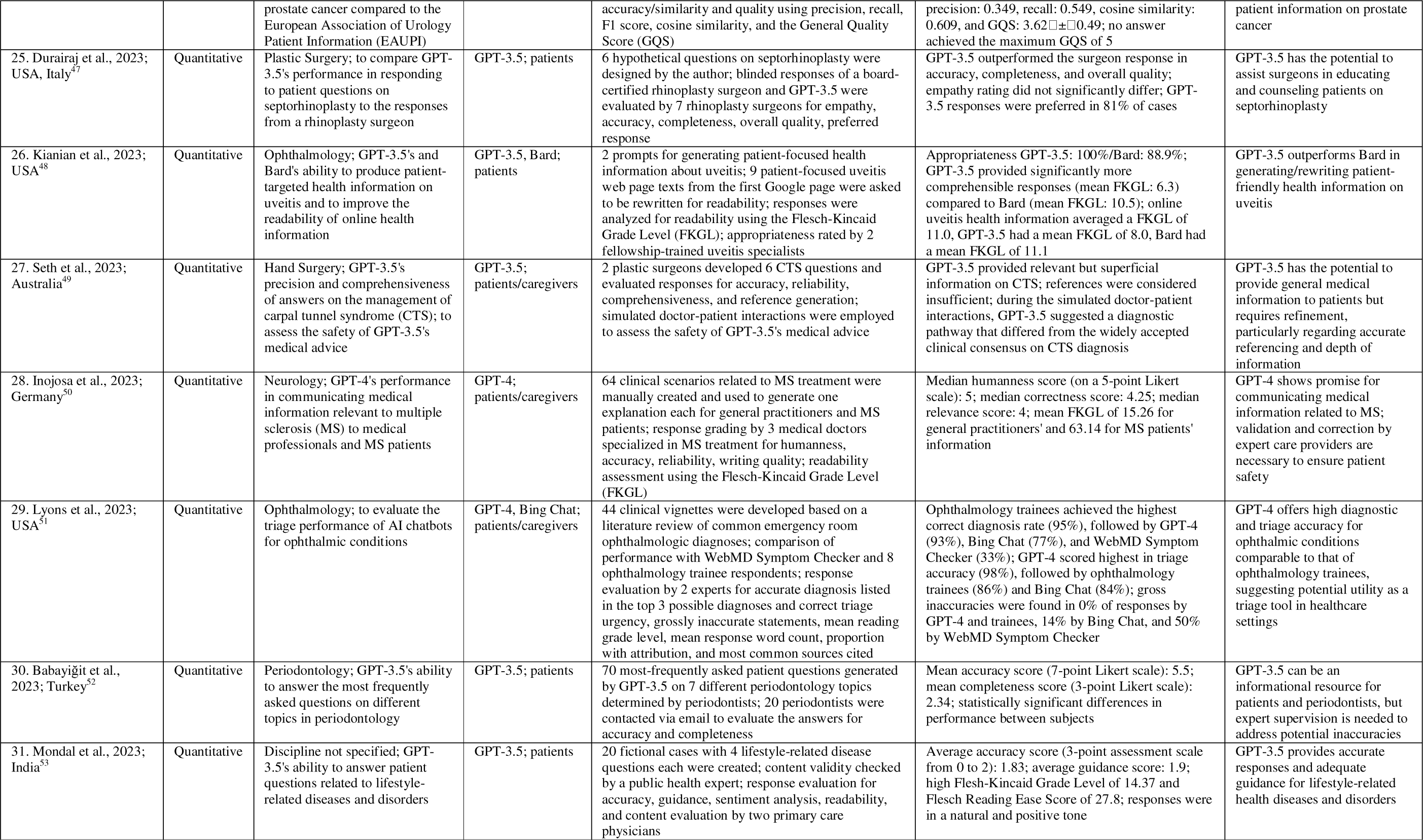
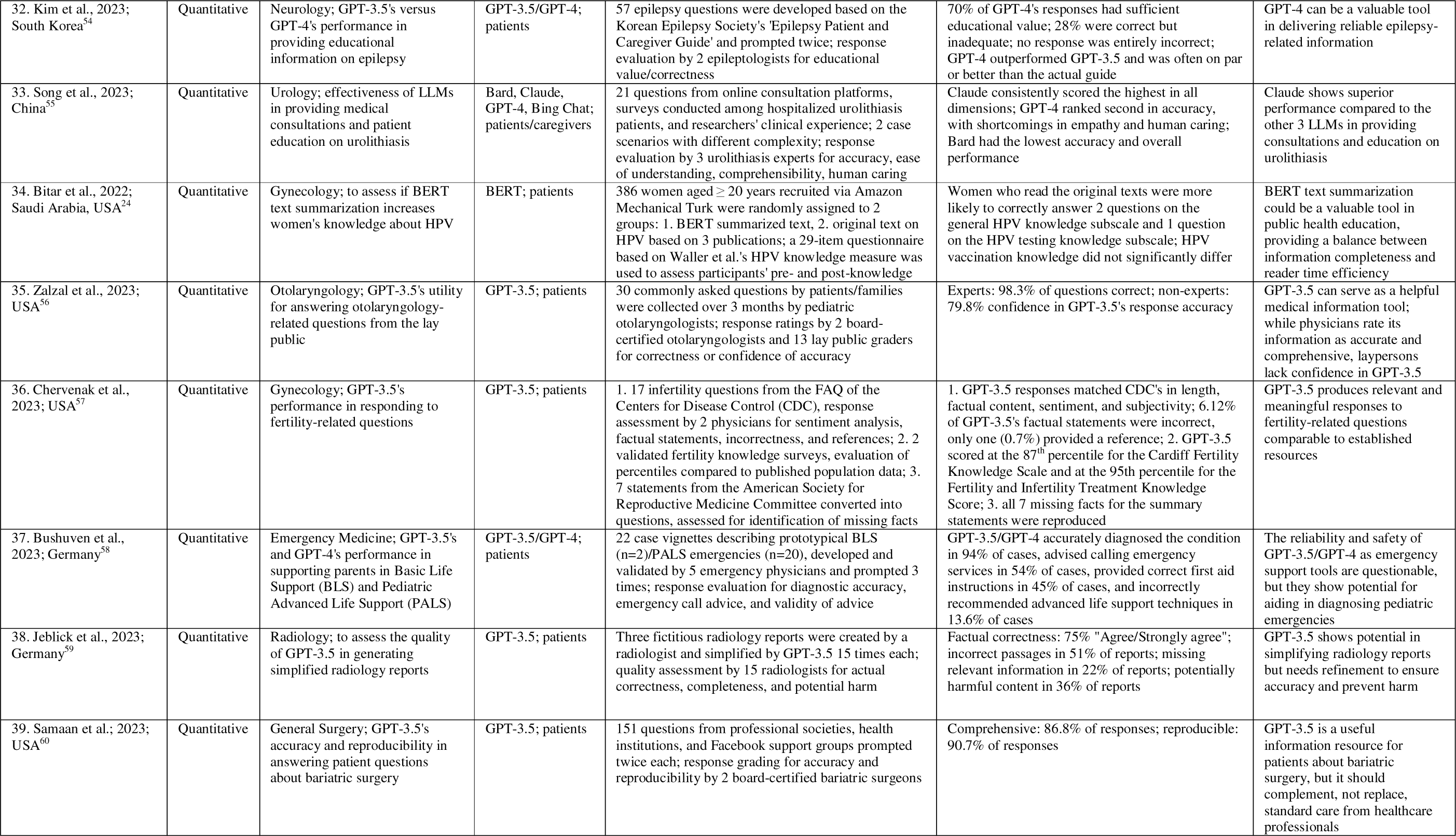
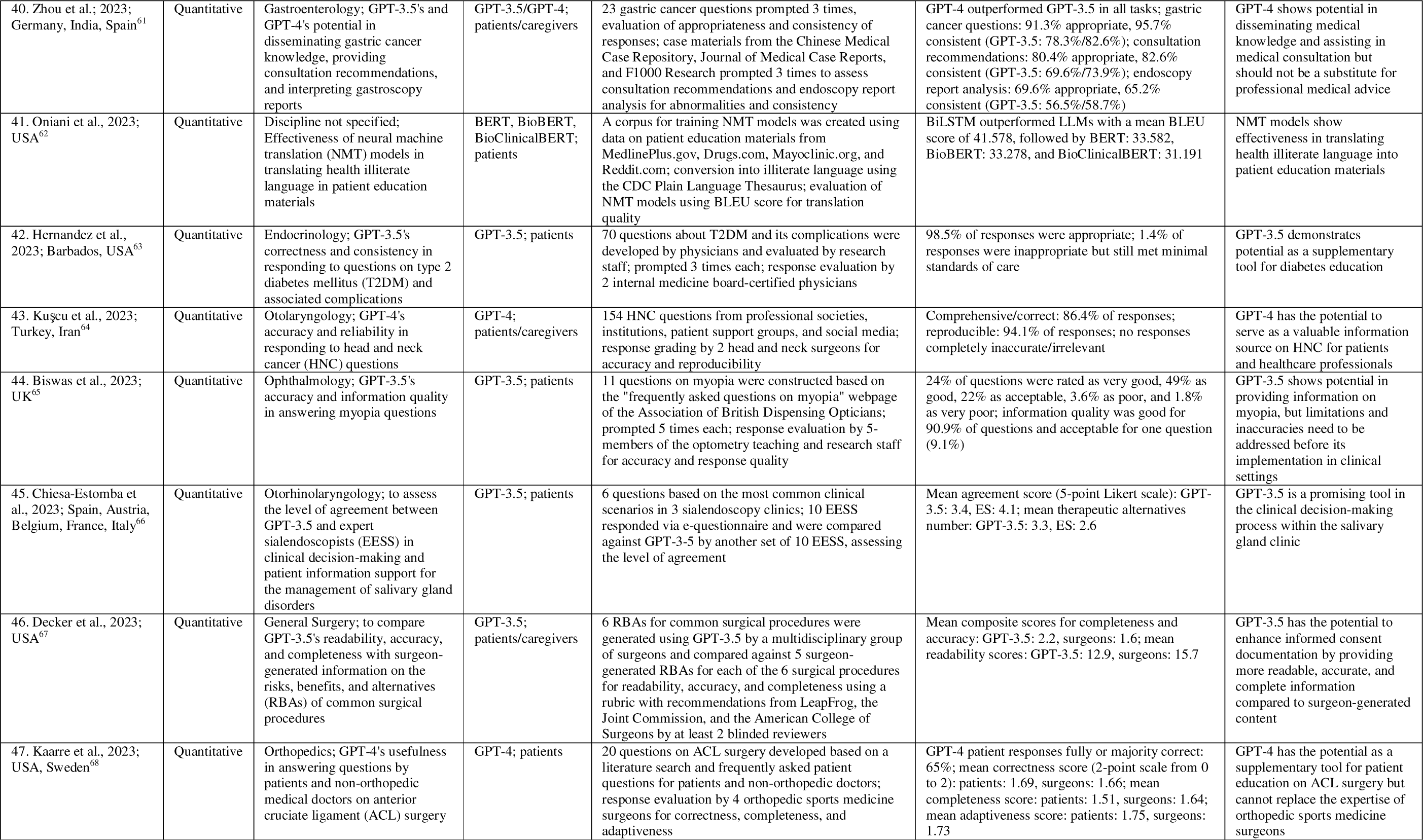
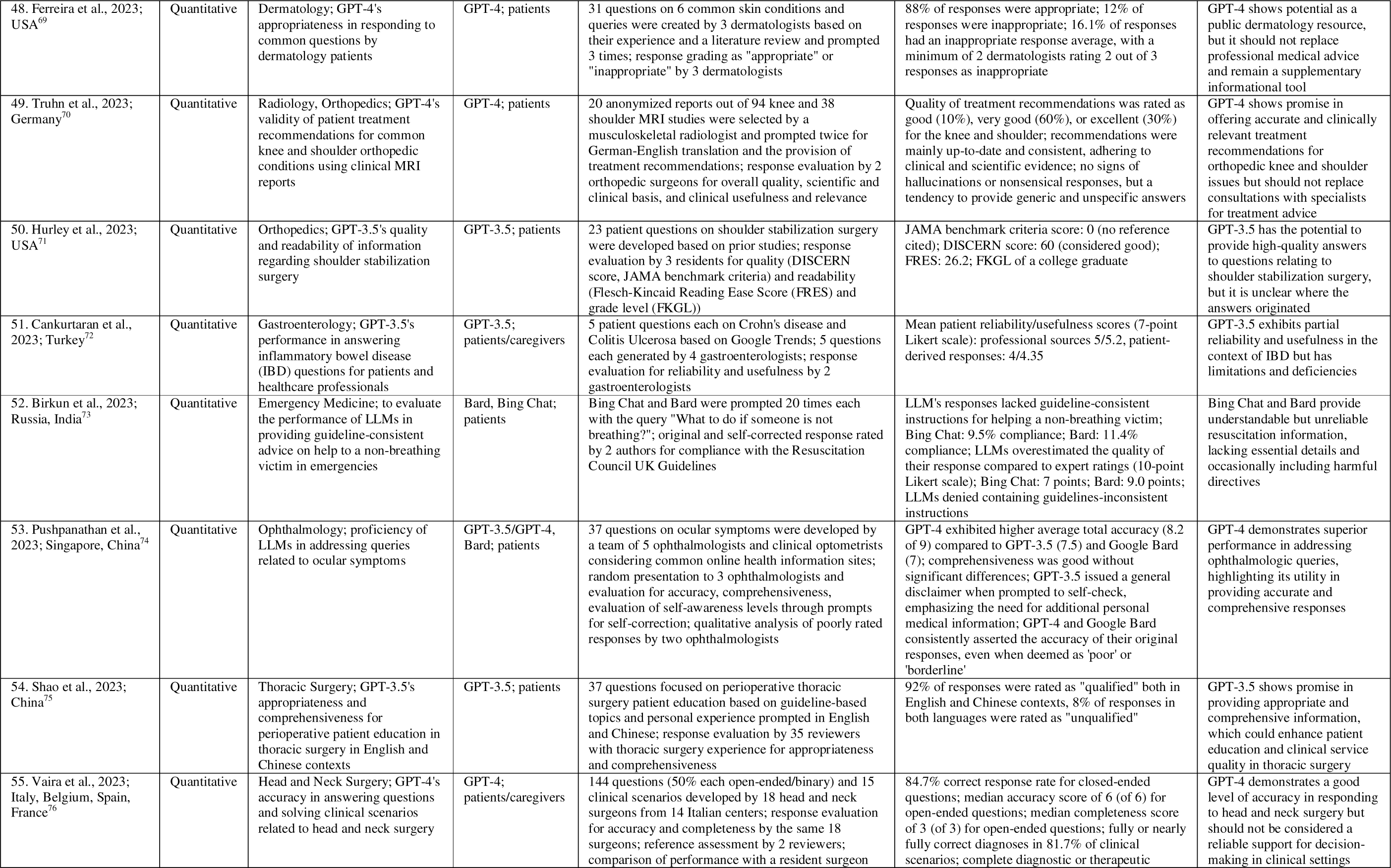
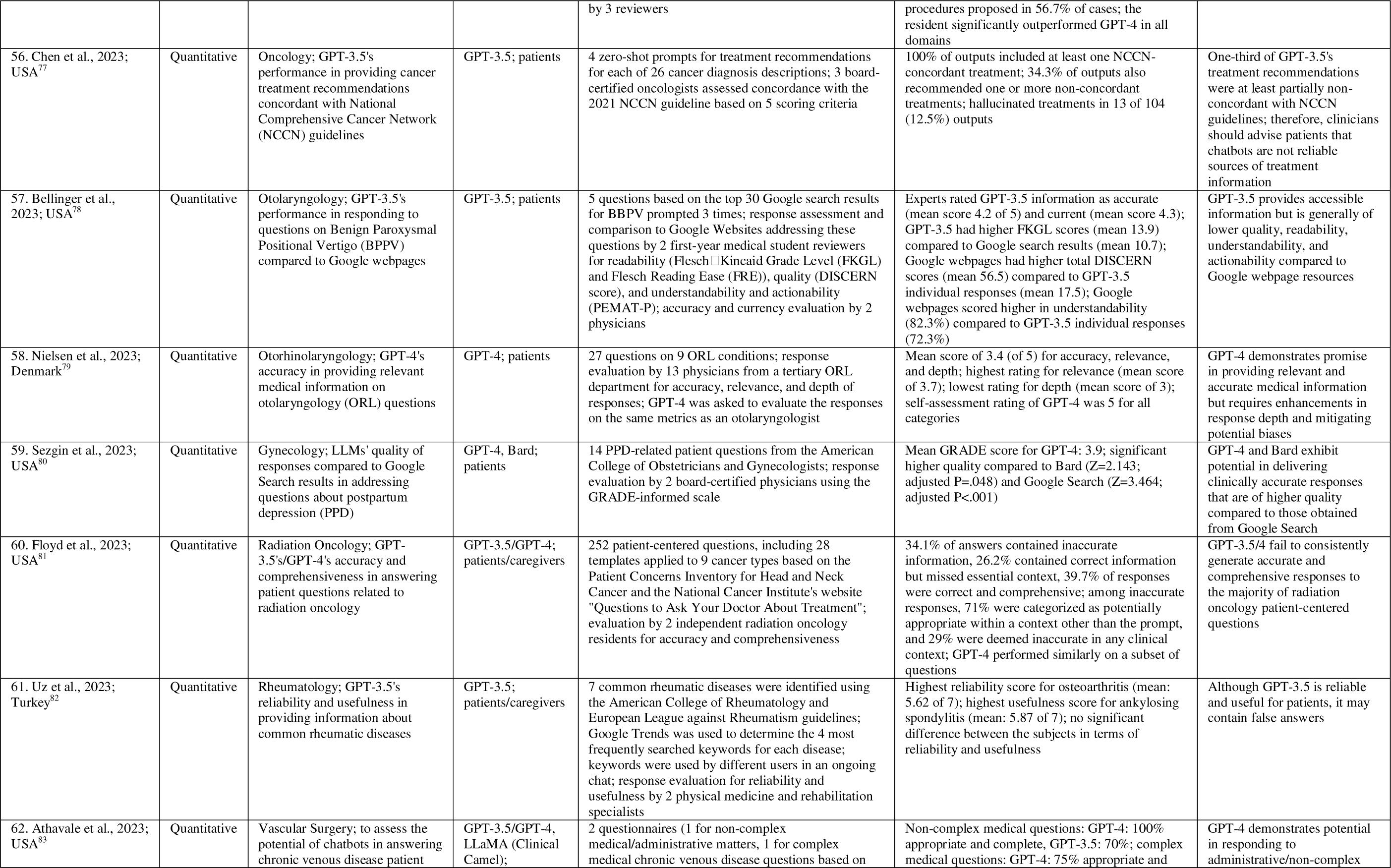
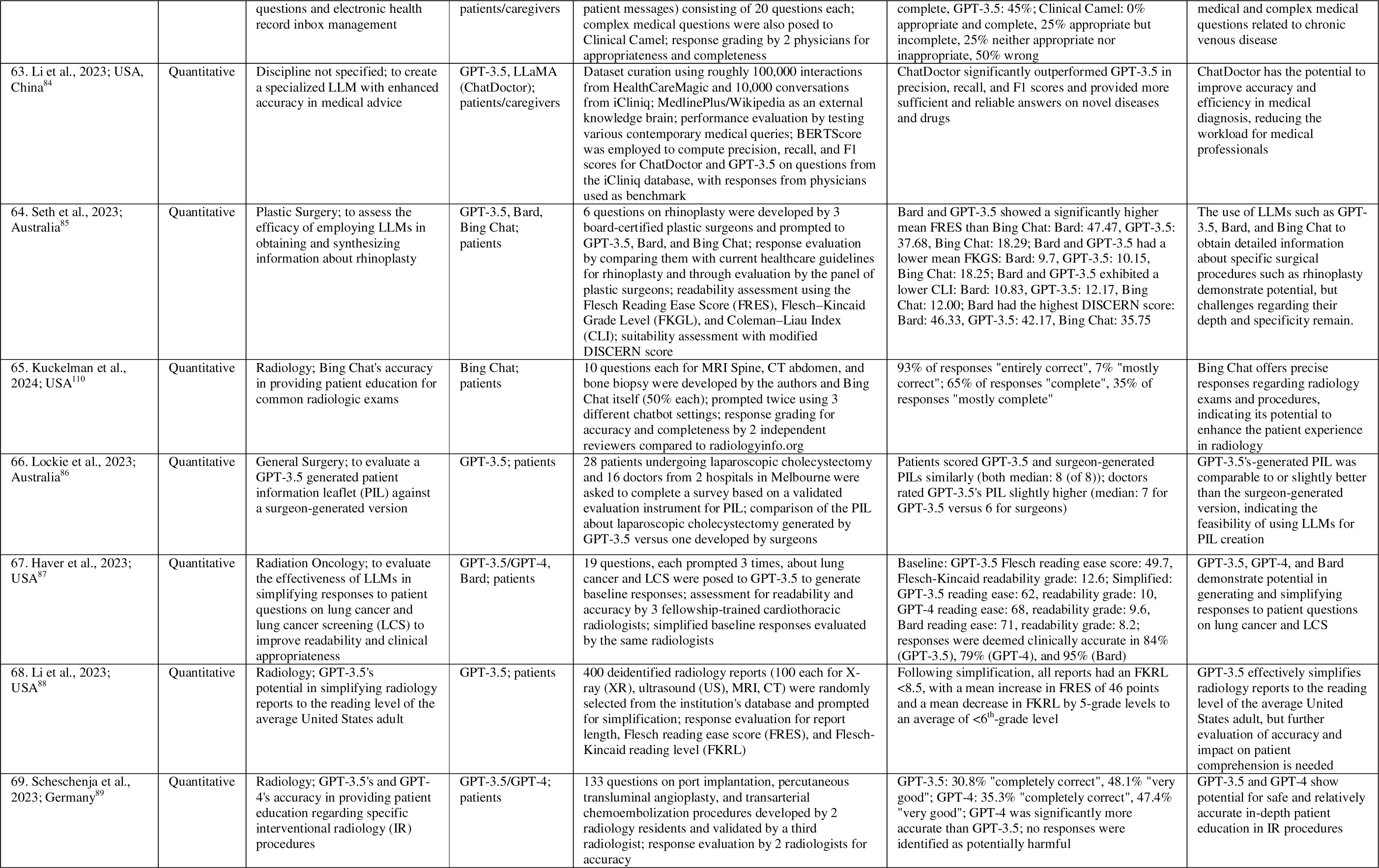
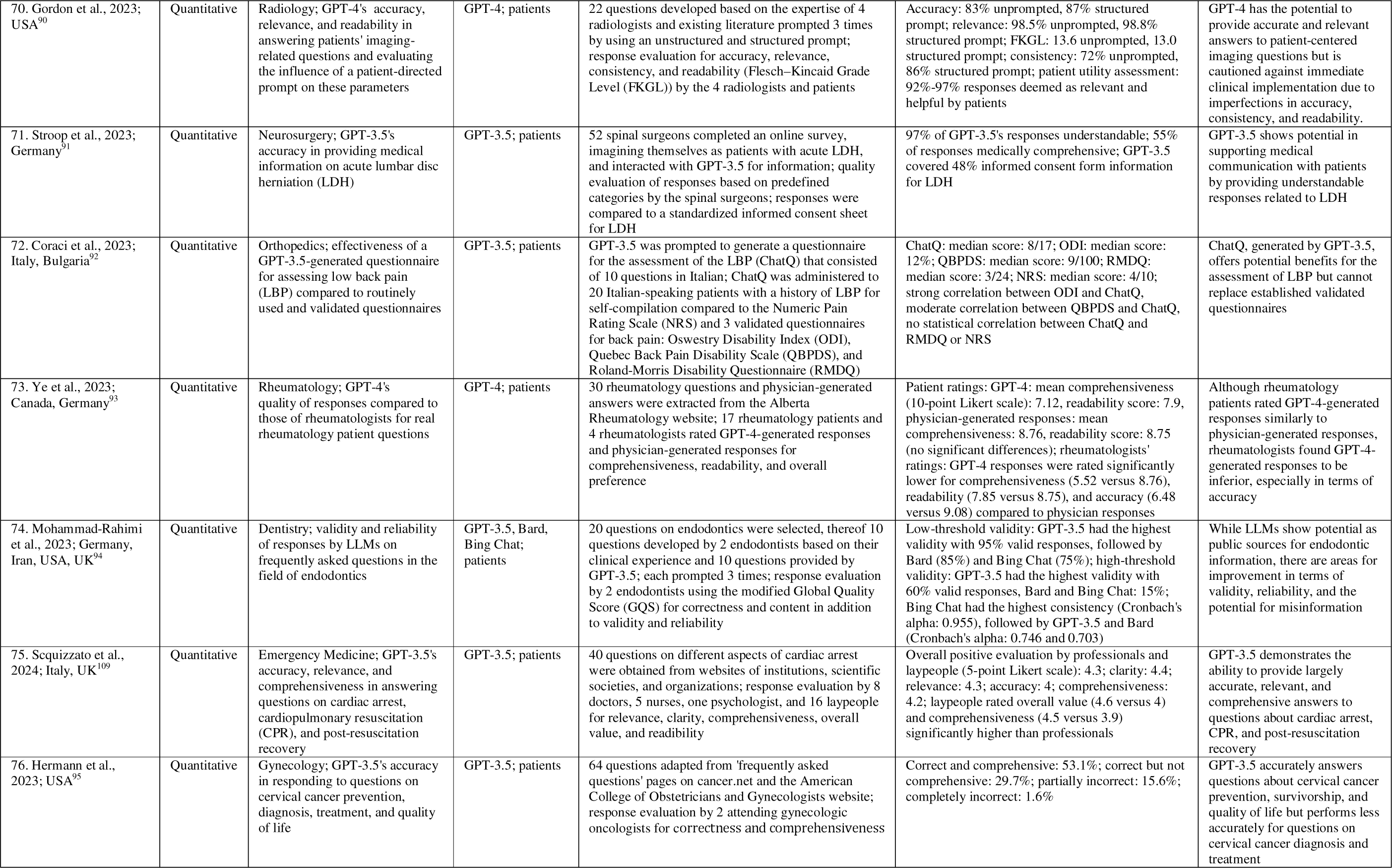
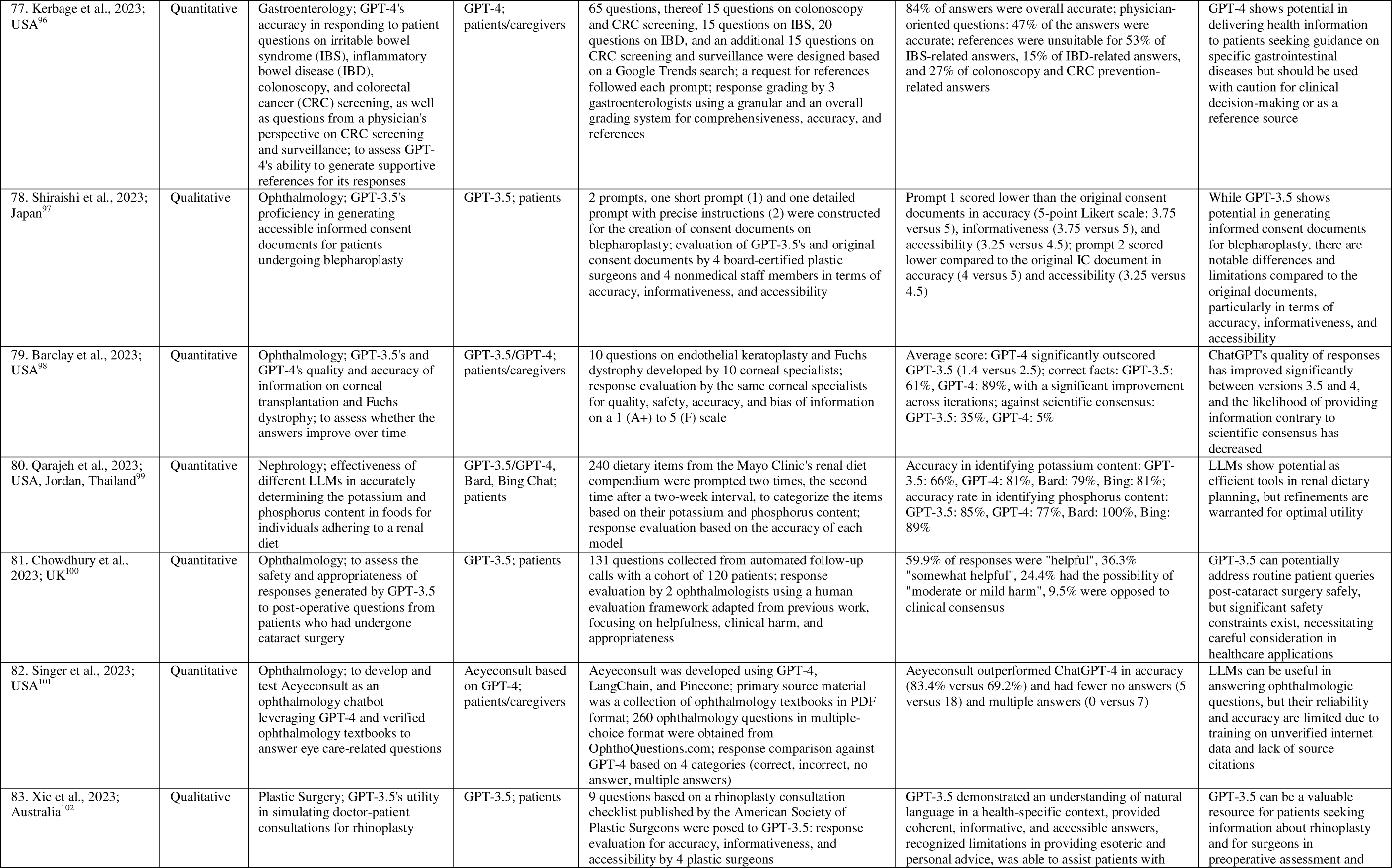
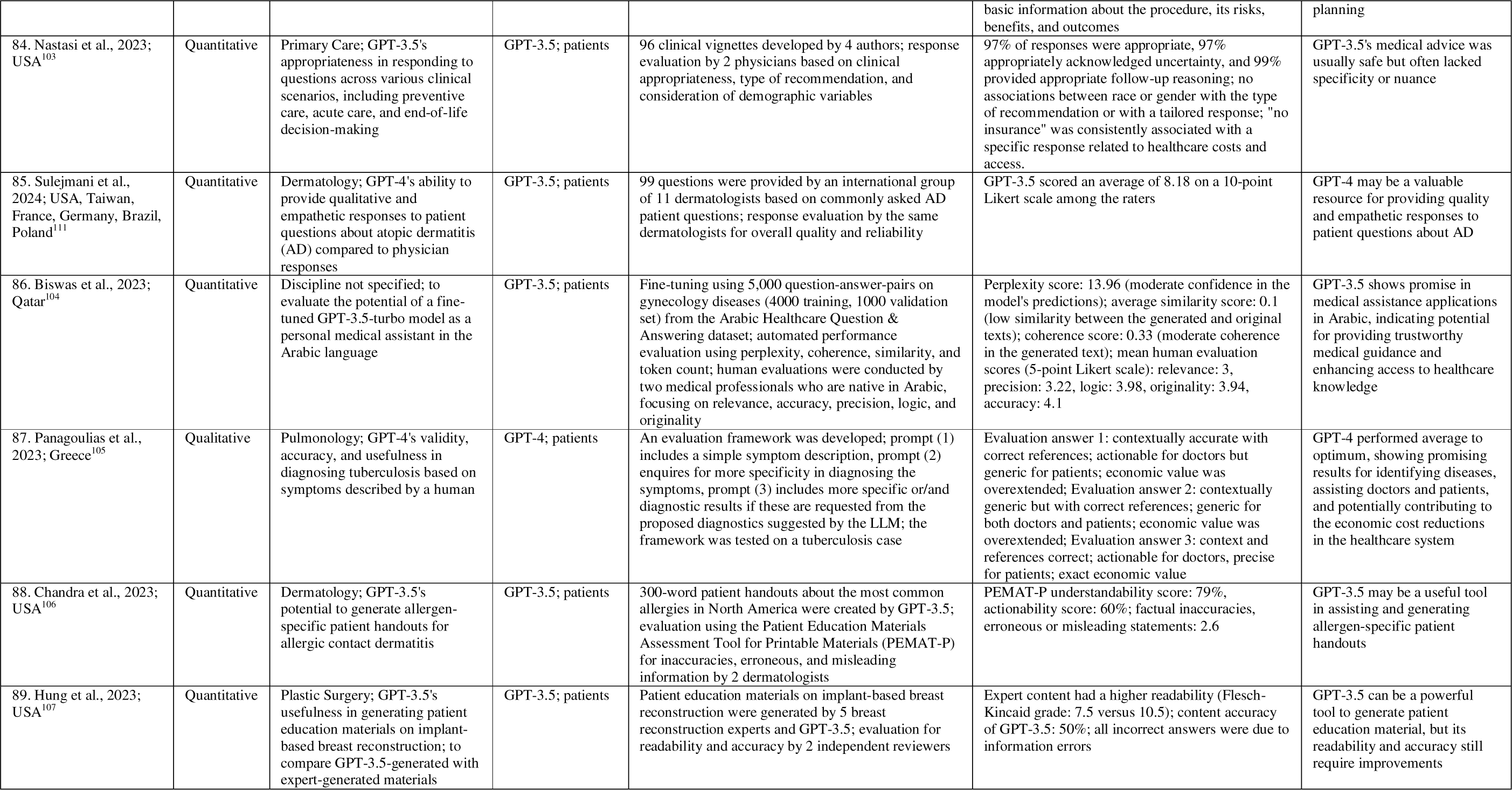
Overview of included studies and corresponding authors, year of publication, affiliation countries of authors, study design, medical specialty, purpose of study, large language model (LLM)/tool examined, target user, evaluation/setting, main outcome, and conclusion.

**Table 2.**
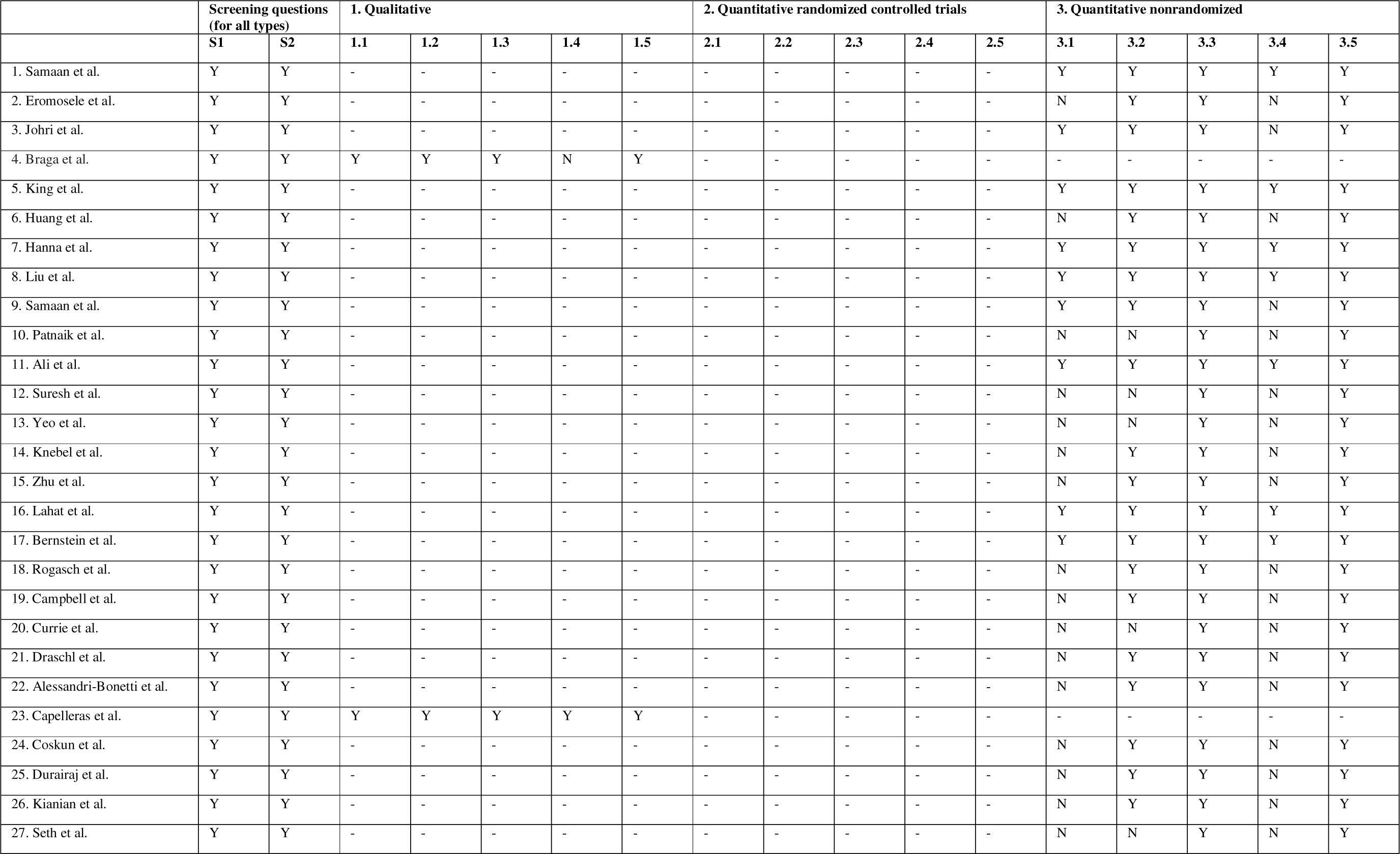
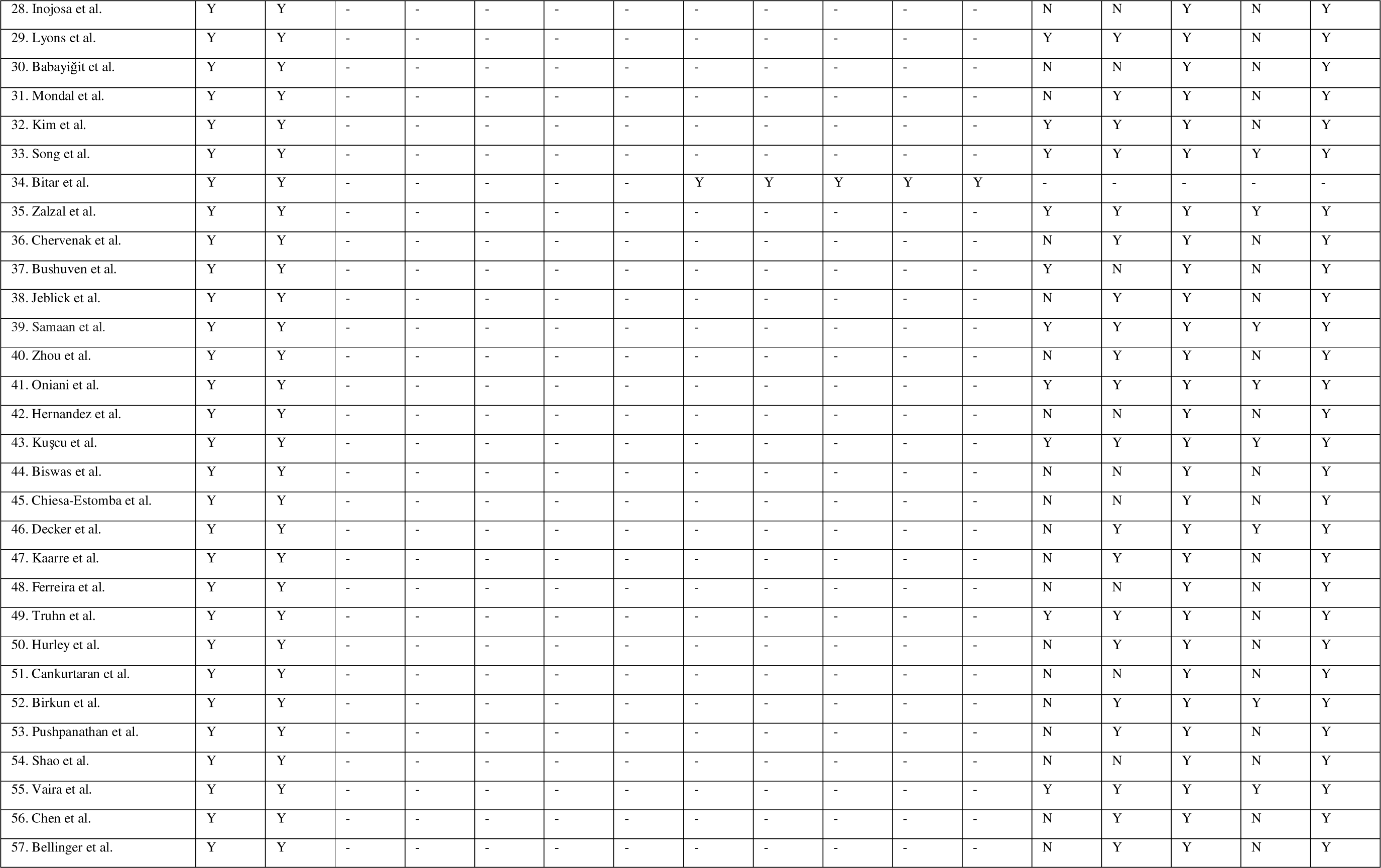
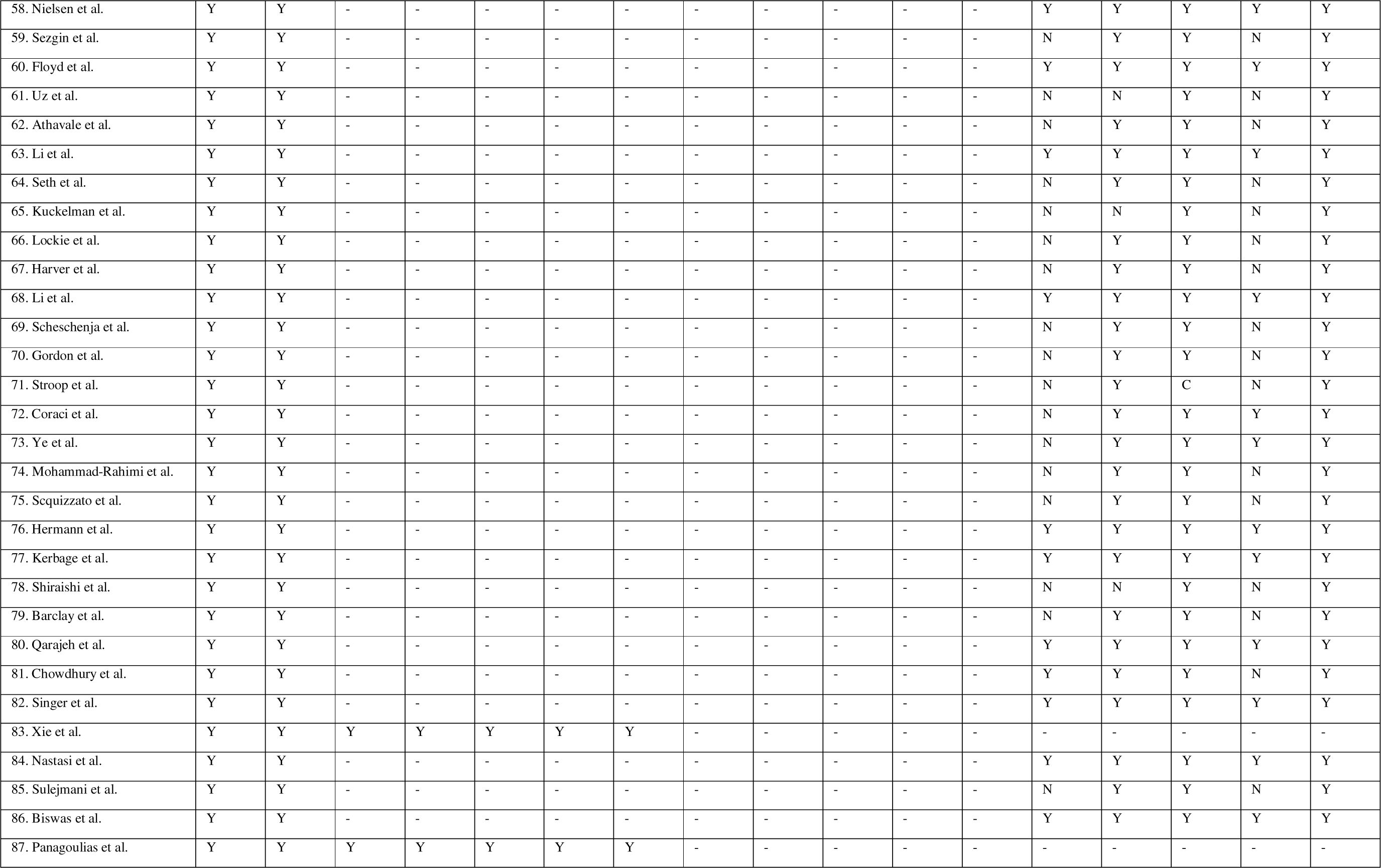
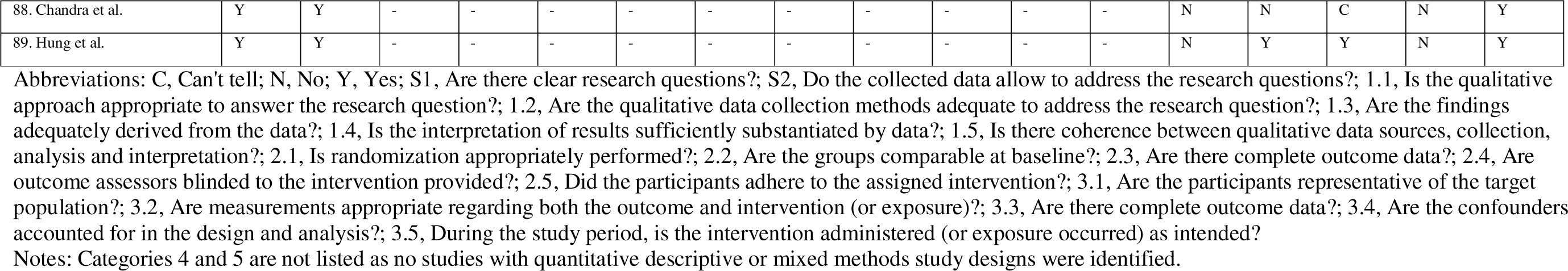
Evaluation of included studies according to the Mixed Methods Appraisal Tool (MMAT) 2018. ^18^

### 3.3 Applications of Large Language Models

An overview of the presence of codes for each study is provided in Supplementary Section 3. The majority of articles investigated the use and feasibility of LLMs as medical chatbots (n=84/89, 94.4%)^13,24–62,64–66,68,69,71–96,98–111^, while fewer reports additionally or exclusively focused on the generation of patient information (n=19/89, 21.4%)^24,31,43,48,49,57,59,62,67,70,79,88–91,97,102,106,107^, including clinical documentation such as informed consent forms (n=5/89, 5.6%)^43,67,91,97,102^ and discharge instructions (n=1/89, 1.1%)^31^, or translation/summarization tasks of medical texts (n=5/89, 5.6%)^24,49,57,79,89^, creation of patient education materials (n=5/89, 5.6%)^48,62,90,106,107^, and simplification of radiology reports (n=2/89, 2.3%)^59,88^. Most reports evaluated LLMs in English (n=88/89, 98.9%)^13,24–103,105–111^, followed by Arabic (n=2/84, 2.3%)^32,104^, Mandarin (n=2/84, 2.3%)^36,75^, and Korean or Spanish (n=1/89, 1.1%, respectively)^75^. The top-five specialties studied were ophthalmology (n=10/89, 11.2%)^37,40,48,51,65,74,97,98,100,101^, gastro-enterology (n=9/89, 10.1%)^25,32,34,36,39,61,62,72,96^, head and neck surgery/otolaryngology (n=8/89, 9%)^35,42,56,64,66,76,78,79^, and radiology^59,70,88–90,110^ or plastic surgery^45,47,49,102,107,108^ (n=6/89, 6.7%, respectively). A schematic illustration of the identified concepts of LLM applications in patient care is shown in Figure 2.

**Figure 2.**
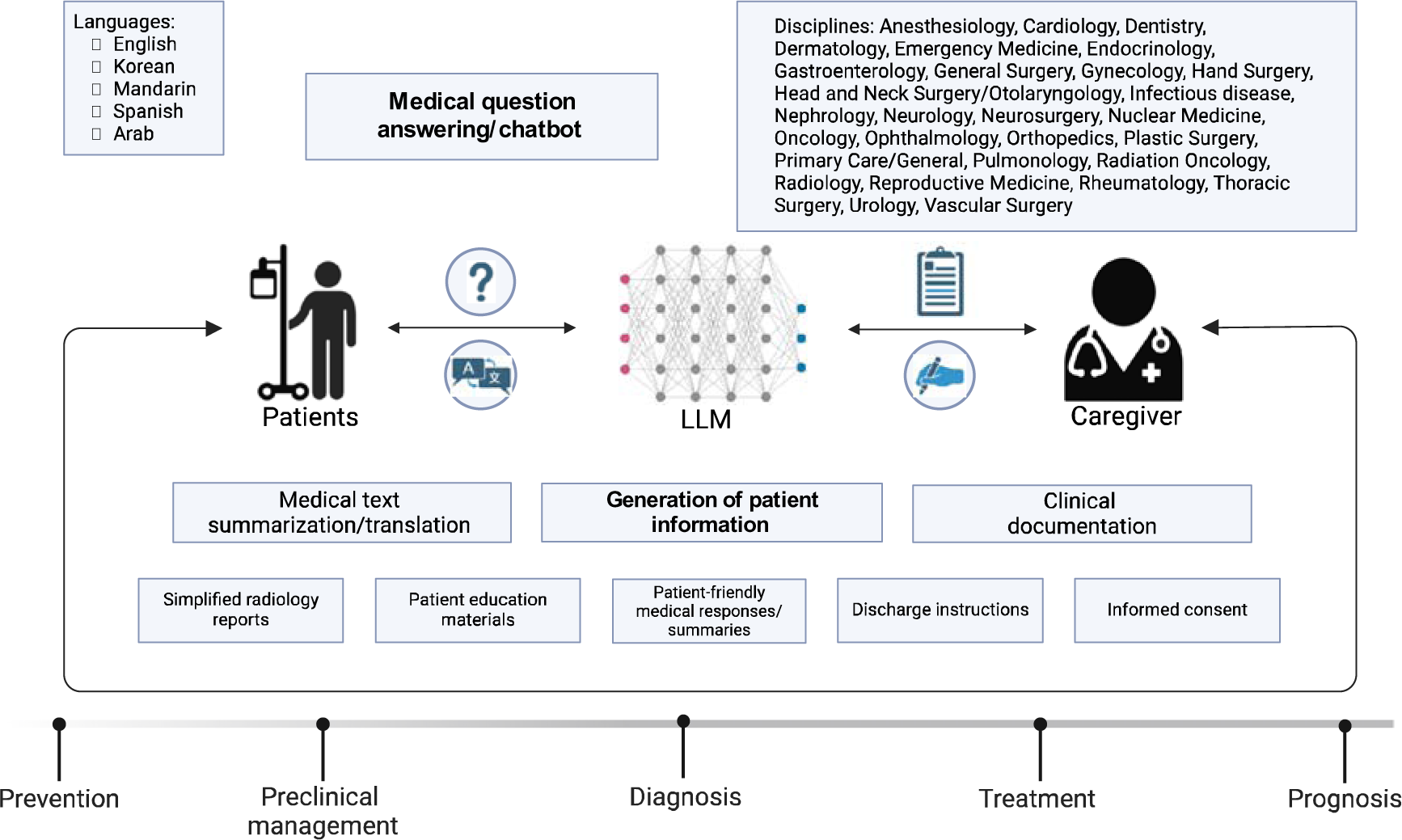
Schematic illustration of the identified concepts for the application of large language models (LLMs) in patient care.

### 3.4 Limitations of Large Language Models

The thematic synthesis of limitations resulted in two main concepts: one related to design limitations and one related to output.

#### 3.4.1 Design limitations

In terms of design limitations, many authors noted the limitation that LLMs are not optimized for medical use (n=46/89, 51.7%)^13,26,28,34,35,37–39,46,49,50,54–59,61,62,65,66,68,70,71,79–81,83–85,88,91,93–98,100–107,109^, including implicit knowledge/lack of clinical context (n=13/89, 14.6%)^28,39,46,66,71,79,81,83–85,98,103^, limitations in clinical reasoning (n=7/89, 7.9%) ^55,84,95,102–105^, limitations in medical image processing/production (n=5/89, 5.6%)^37,55,91,106,107^, and misunderstanding of medical information and terms by the model (n=7/89, 7.9%)^28,38,39,59,62,65,97^. In addition, data-related limitations were identified, including limited access to data on the internet (n=22/89, 24.7%)^38,39,41,43,54–57,59,60,64,76,79,82–84,88,91,94,96,104,109^, the undisclosed origin of training data (n=36/89, 40.5%)^25,26,29,30,32,34,36,37,40,46,47,50,51,53–60,64,65,70,71,76,82,83,91,94–96,101,105,109^, limitations in providing, evaluating, and validating references (n=20/89, 22.5%)^45,49,54–57,65,71,73,76,80,83,85,91,94,96,98,101,103,105^, and storage/processing of sensitive health information (n=8/89, 9%)^13,34,46,55,62,76,83,109^. Further second-order concepts included black-box algorithms, i.e., non-explainable AI (n=12/89, 13.5%)^27,36,55,57,65,73,76,83,91,94,103,105^, limited engagement and dialogue capabilities (n=10/89)^13,27,28,37,38,51,56,66,95,103^, and the inability of self-validation and correction (n=4/89, 4.5%)^61,73,74,107^.

#### 3.4.2 Output limitations

The evaluation of limitations in output data yielded 7 second-order codes concerning the non-reproducibility (n=38/89, 42.7%)^28,29,34,38,39,41,43,45,46,49,54–61,64,65,71–73,76,80,82,83,85,90,91,94,96,98,99,101,103–105^, non-comprehensiveness (n=78/89, 87.6%)^13,25,26,28–30,32–44,46,48–62,64,65,67–79,81–98,100,102–107,109–111^, incorrectness (n=78/89, 87.6%)^13,25–44,46,49–52,54–62,64–66,69–79,81–85,87–107,109–111^, (un-)safeness (n=39/89, 43.8%)^28,30,35,37,39,40,42–44,46,50,51,57–60,62,64,65,69,70,73,74,76,78–80,82,84,85,91,94,95,98–100,105,106,109^, bias (n=6/89, 6.7%)^26,32,34,36,66,103^, and the dependence of the quality of output on the prompt-/input provided (n=27/89, 30.3%)^26–28,34,38,41,44,46,51,52,56,68–72,74,76,78,79,81–83,90,94,95,100,101^ or the environment (n=16/89, 18%)^13,34,46,49–51,54,58,60,72,73,88,90,93,97,109^.

For non-reproducibility, key concepts included the non-deterministic nature of the output, e.g., due to inconsistent results across multiple iterations (n=34/89, 38.2%)^28,29,34,38,39,41,43,46,58–61,72,76,82,90,94,98,99,101,103,104^ and the inability to provide reliable references (n=20/89, 22.5%)^45,49,54–57,65,71,73,76,80,83,85,91,94,96,98,101,103,105^. Non-comprehensiveness included nine concepts related to generic/non-personalized output (n=34/89, 38.2%)^13,28,30,34,37,38,41,43,49,51,56,57,59,61,65,70,77,79,81,84–86,90,94,95,100,102–107,110^, incompleteness of output (n=68/89, 76.4%)^13,25,26,28–30,32,34–39,41–44,46,49–52,55–62,64,65,67–69,72–77,79,81–86,89–98,100,102–107,109–111^, provision of information that is not standard of care (n=24/89, 27%)^28,40,43,46,49,50,54,57,58,65,69,72,73,77,78,81,85,91,94,98,100,103,107,111^ and/or outdated (n=12/89, 13.5%)^13,25,32,34,38,41,43,44,49,54,83,84^, and production of oversimplified (n=10/89, 11.2%)^38,46,49,54,59,79,84,85,103^, superfluous (n=16/89, 18%)^13,28,34,38,46,62,72,79,86,90,94,97,100,106,107^, overcautious (n=7/89, 7.9%)^13,28,37,51,70,103,110^, overempathic (n=1/89, 1.1%)^13^, or output with inappropriate complexity/reading level for patients (n=22/89, 24.7%)^13,34,42,48,50,51,53,55,56,67,71,78,79,85,87,88,90,93,106,107,109,110^. For incorrectness, we identified 6 key concepts. Some of the incorrect information could be attributed to what is commonly known as hallucination (n=38/89, 42.7%)^25,28,32,33,35–38,40–44,49–51,57–60,65,73,74,76,77,81,83,85,91,94,96–98,100,103,106,107,109^, i.e., the creation of entirely fictitious or false information that has no basis in the input provided or in reality (e.g., “You may be asked to avoid eating or drinking for a few hours before the scan” for a bone scan). However, numerous instances of misinformation were more appropriately classified under alternative concepts of the original psychiatric analogy, as described in detail by Currie et al.^43,112,113^ These include illusion (n=12/89, 13.5%)^28,36,38,43,57,59,77,78,85,88,94,105^, which is characterized by the generation of deceptive perceptions or the distortion of information by conflating similar but separate concepts (e.g., suggesting that MRI-type sounds might be experienced during standard nuclear medicine imaging), delirium (n=34/89, 38.2%)^13,26,28,30,37,43,50,58,59,61,65,70,72–75,77,79,81–85,90–92,94,95,98,102,103,107,109,110^, which indicates significant gaps in vital information, resulting in a fragmented or confused understanding of a subject (e.g., omission of crucial information about caffeine cessation for stress myocardial perfusion scans), extrapolation (n=11/89, 12.4%)^43,59,65,78,81,91,94,106,107,110^, which involves applying general knowledge or patterns to specific situations where they are inapplicable (e.g., advice about injection-site discomfort that is more typical of CT contrast administration), delusion (n=14/89, 15.7%)^28,30,43,50,59,65,69,73,74,78,81,94,103,111^, a fixed, false beliefs despite contradictory evidence (e.g., inaccurate waiting times for the thyroid scan), and confabulation (n=18/89, 20.2%)^25,28,36–38,40,46,59,62,65,71,77–79,94,103,107^, i.e., filling in memory or knowledge gaps with plausible but invented information (e.g., “You should drink plenty of fluids to help flush the radioactive material from your body” for a biliary system–excreted radiopharmaceutical).

Many studies rated the generated output as unsafe, including misleading (n=34/89, 38.2%)^28,30,35,43,44,46,50,51,57–60,62,64,65,69,73,74,76,78–80,82,84,85,94,95,98–100,105,106,109^ or even harmful content (n=26/89, 29.2%)^28,30,37,39,40,42,43,50,51,58–60,70,73,74,76,79,84,85,91,94,95,98–100,109^. A minority of reports identified biases in the output, which were related to language (n=2/89, 2.3%)^32,36^, insurance status^103^, underserved racial groups^26^, or underrepresented procedures^34^ (n=1/89, 1.1%, each). Finally, many authors suggested that performance was related to the prompting/input provided or the environment, i.e., depending on the evidence (n=7/89, 7.9%)^52,68,69,71,81,82,95^, complexity (n=11/89, 12.4%)^28,34,44,46,70,74,76,79,94,102^, specificity (n=13/89, 14.6%)^27,38,41,56,70,72,74,76,78,81,95,100,101^, quantity (n=3/89, 3.4%)^26,52,74^ of the input, type of conversation (n=3/89, 3.4%)^27,51,90^, or the appropriateness of the output related to the target group (n=9/89, 10.1%)^46,49,51,54,72,90,93,97,109^, provider/organization (n=4/89, 4.5%)^13,50,60,88^, and local/national medical resources (n=5/89, 5.6%)^34,50,58,60,73^. Figure 3 illustrates the hierarchical tree structure and quantity of the codes derived from the thematic synthesis of limitations.

**Figure 3.**
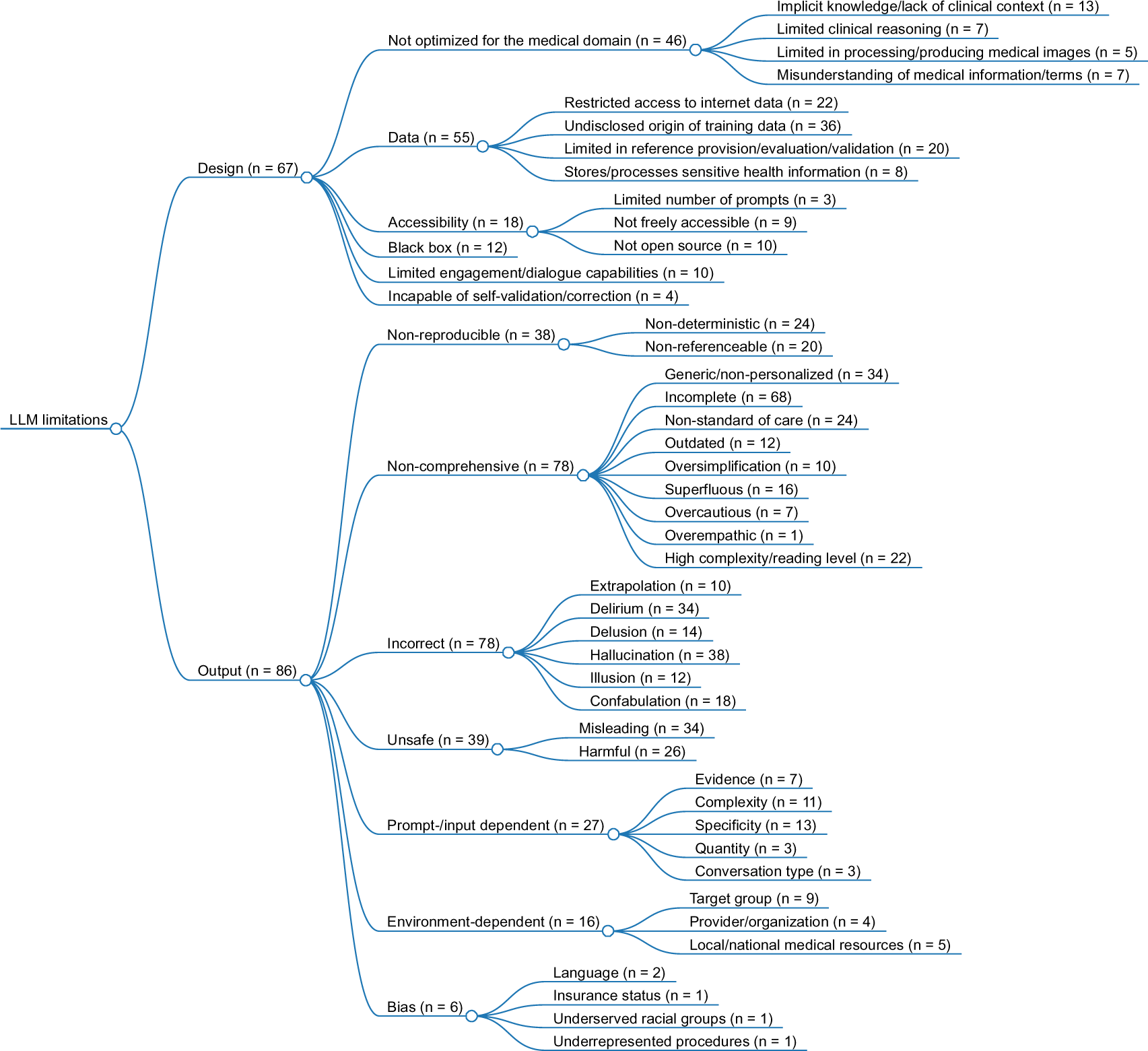
Illustration of the hierarchical tree structure for the thematic synthesis of large language model (LLM) limitations in patient care, including the presence of codes for each concept.

## 4. Discussion

In this systematic review, we synthesized the current applications and limitations of LLMs in patient care, incorporating a broad analysis across 29 medical specialties and highlighting key limitations in LLM design and output, providing a comprehensive framework and taxonomy for their future implementation and evaluation in healthcare settings.

Most articles examined the use of LLMs based on the GPT-3.5 or GPT-4 architecture for answering medical questions, followed by the generation of patient information, including medical text summarization or translation and clinical documentation. The conceptual synthesis of LLM limitations revealed two key concepts: the first related to design, including 6 second-order and 12 third-order codes, and the second related to output, including 9 second-order and 32 third-order codes.

Although many LLMs have been developed specifically for the biomedical domain in recent years, we found that ChatGPT has been a disruptor in the medical literature on LLMs, with GPT-3.5 and GPT-4 accounting for almost 80% of the LLMs examined in this systematic review. While it was not possible to conduct a meta-analysis of the performance on medical tasks, many authors provided a positive outlook towards the integration of LLMs into clinical practice. However, the use of proprietary models such as ChatGPT in the biomedical field raises concerns because the limited access to the underlying algorithms, training data, and data processing and storage mechanisms makes them untransparent and, thus, significantly limits their applicability in healthcare.^114^ Furthermore, the integration of proprietary models into patient care applications makes one susceptible to performance changes associated with model updates, which may break existing functionalities and lead to harmful outcomes for patients. Therefore, especially in the biomedical field, open-source models such as BioMistral may offer a viable solution.^6^ Given the limited number of articles on open-source LLMs in our review, we strongly encourage future studies investigating the applicability of open-source LLMs in patient care. We identified several key limitations regarding the design and output. Not surprisingly, many reports noted the limitation that the LLMs studied were not optimized for the medical domain. One possible solution to this limitation may be to provide medical knowledge during inference using RAG.^115^ However, even when trained for general purposes, ChatGPT has previously been shown to pass the United States Medical Licensing Examination (USMLE), the German State Examination in Medicine, or even a radiology board-style examination without images.^116–119^ Although outperformed on specific tasks by specialized medical LLMs, such as Google’s MedPaLM-2, this suggests that general-purpose LLMs can comprehend complex medical literature and case scenarios to a degree that meets professional standards.^120^ Furthermore, given the large amounts of data on which proprietary models such as ChatGPT are trained, it is not unlikely that they have been exposed to more medical data overall than smaller specialized models despite being generalist models.

It should also be noted that passing these exams does not equate to the practical competence required of a healthcare provider.^121^ In addition, reliance on exam-based assessments carries a significant risk of bias. For example, if the exam questions or similar variants are publicly available and, thus, may be present in the training data, the LLM does not demonstrate any knowledge outside of training data memorization.^122^ In fact, these types of tests can be misleading in estimating the model’s true abilities in terms of comprehension or analytical skills. Many studies have reported limitations in the output related to comprehensiveness, safety, correctness, reproducibility, and dependence of the output on the input/prompt and environment. Specifically, for correctness, we followed the taxonomy of Currie et al. to classify incorrect outputs more precisely into illusions, delusions, delirium, confabulation, and extrapolation, thus proposing a framework for a more precise and structured error classification to improve the characterization of incorrect outputs and enabling more detailed performance comparisons with other research.^43,112,113^ On the other hand, a minority of studies have identified biases, for example, reflecting the unequal representation of certain content or the biases inherent in human-generated text in the training data.^123^ This may indicate that the implemented safeguards are effective. However, not much is known about the technology and developer policies of proprietary LLMs, and previous work has shown that automated jailbreak generation is possible across various commercial LLM chatbots.^124^ This also mirrors our concept of data-related limitations, particularly regarding the handling of sensitive health information. Together with the limited transparency about the origin of the training data and the unexplainable and non-deterministic nature of the output, this raises a key question when applying LLMs to the medical domain: how can we entrust our patients to LLMs if they are neither reliable nor transparent? Given that models like ChatGPT are already publicly accessible and widely used, patients may already refer to them for medical questions in much the same way they use Google Search, making concerns about their early adoption somewhat academic.^125^

In addition, low health literacy due to the identified limitations in comprehensiveness, including the generation of content with high complexity and an inappropriate reading level, which was above the 6th-grade level recommended by the American Medical Association (AMA) in almost all studies analyzed, may further limit their utility for patient information.^126^ Overall, this can lead to results that are misleading and harmful, as described in many of the reports in our review. In addition to advances in the development of LLMs and the focus on open source, it will therefore be necessary to develop and implement a well-validated scale to determine the quality and safety of LLM outputs in medical practice, such as the recent effort made to adopt the widely recognized Physician Documentation Quality Instrument (PDQI-9) for the assessment of AI transcripts and clinical summaries.^127^

Finally, the implementation of regulatory mandates like the forthcoming European Union AI Act and the associated challenges faced by generative AI and LLMs, for example, in terms of training data transparency and validation of non-deterministic output, will show which approaches the companies will take to bring these models into compliance with the law. How the notified bodies interpret and enforce the law in practice will likely be decisive for the further development of LLMs in the biomedical sector.^128^

### 4.1 Limitations

Our study has limitations. First, our review focused on LLM applications and limitations in patient care, thus excluding research directed at clinicians only. Future studies may extend our synthesis approach to LLM applications that explicitly focus on healthcare professionals. Second, there is a risk that potentially eligible studies were not included in our analysis if they were not present in the 5 databases reviewed or were not available in English. However, we screened nearly 3,000 articles in total and systematically analyzed 89 articles, providing a comprehensive overview of the current state of LLMs in patient care, even if some articles could have been missed. Third, the rapid development and advancement of LLMs make it difficult to keep this systematic review up to date. For example, Gemini 1.5 Pro was published in February 2024, and corresponding articles are not included in this review, which synthesized articles from 2022 to 2023. Continued updates will be essential to monitor emerging areas and limitations in this rapidly evolving field.

## 5. Conclusion

In conclusion, this review provides a systematic overview of current LLM applications and limitations in patient care. Our conceptual synthesis provides a structured taxonomy that may lay the groundwork for both the implementation and critical evaluation of LLMs in healthcare settings.

## 6. Declarations

## Supporting information

Supplementary Sections 1-3

## 6.1 Acknowledgements

This research is funded by the European Union (101079894). Views and opinions expressed are however those of the authors only and do not necessarily reflect those of the European Union or European Commission. Neither the European Union nor the granting authority can be held responsible for them. The funding had no role in the study design, data collection and analysis, manuscript preparation, or decision to publish.

## 6.3 Author contributions

Conceptualization: FB, LCA, KKB; Project administration: FB; Resources: FB, LCA, KKB; Software: FB, LCA, KKB; Data curation: FB, LH, CR; Formal analysis: FB, LH, CR, LCA, KKB; Investigation: FB, LH, CR, LCA, KKB; Methodology: FB; Supervision: FB, LCA, KKB; Validation: FB, LH, CR, EHCvD, RK, EOP, MRM, LS, MH, JNK, DT, RC, LCA, KKB; Visualization: FB, LCA; Writing – original draft preparation: FB, LH, LCA, KKB; Writing – review & editing: FB, LH, CR, EHCvD, RK, EOP, MRM, LS, MH, JNK, DT, RC, LCA, KKB.

## 6.4 Data availability

All data generated or analyzed during this study are included in this published article and its supplementary information files.

