## Supplementary Sections 1-3 for "Systematic Review of Large Language Models for Patient Care: Current Applications and Challenges"

### Supplemental material

#### Section 1. Database search strategy

**1. Web of Science**
TS=("large language model" OR "LLM" OR "GPT" OR "BERT" OR "transformer model" OR "generative model" OR "generative AI" OR "generative artificial intelligence" OR "generative pre-trained transformer" OR "bidirectional encoder representations from transformers" OR "AI language model") AND TS=(medical practice OR healthcare OR clinical practice OR medicine OR medical treatment OR patient) AND PY=(2022-2023) AND LA=(English) NOT DT=(Review)

**2. PubMed**
("large language model" OR "LLM" OR "GPT" OR "BERT" OR "transformer model" OR "generative model" OR "generative AI" OR "generative artificial intelligence" OR "generative pre-trained transformer" OR "bidirectional encoder representations from transformers" OR "AI language model") AND (medical practice OR healthcare OR clinical practice OR medicine OR medical treatment OR patient) NOT Review[Publication Type] 2022:2023 [dp] English[la]

**3. Embase and Embase Classic**
*Limit to (english language and yr="2022 - 2023")*
("large language model" OR "LLM" OR "GPT" OR "BERT" OR "transformer model" OR "generative model" OR "generative AI" OR "generative artificial intelligence" OR "generative pre-trained transformer" OR "bidirectional encoder representations from transformers" OR "AI language model").mp. AND (medical practice OR healthcare OR clinical practice OR medicine OR medical treatment OR patient).mp. NOT "Review".pt.

**4. ACM Digital Library**
[[Title: "large language model"] OR [Abstract: "large language model"] OR [Title: "llm"] OR [Abstract: "llm"] OR [Title: "gpt"] OR [Abstract: "gpt"] OR [Title: "bert"] OR [Abstract: "bert"] OR [Title: "transformer model"] OR [Abstract: "transformer model"] OR [Title: "generative model"] OR [Abstract: "generative model"] OR [Title: "generative artificial intelligence"] OR [Abstract: "generative artificial intelligence"] OR [Title: "generative pre-trained transformer"] OR [Abstract: "generative pre-trained transformer"] OR [Title: "bidirectional encoder representations from transformers"] OR [Abstract: "bidirectional encoder representations from transformers"] OR [Title: "ai language model"] OR [Abstract: "ai language model"]] AND [[Title: medical practice] OR [Abstract: medical practice] OR [Title: healthcare] OR [Abstract: healthcare] OR [Title: clinical practice] OR [Abstract: clinical practice] OR [Title: medicine] OR [Abstract: medicine] OR [Title: medical treatment] OR [Abstract: medical treatment] OR [Title: patient] OR [Abstract: patient]] AND [E-Publication Date: (01/01/2022 TO 12/31/2023)]

**5. IEEE Xplore**
*Year range 2022-2023*
(("large language model" OR "LLM" OR "GPT" OR "BERT" OR "transformer model" OR "generative model" OR "generative AI" OR "generative artificial intelligence" OR "generative pre-trained transformer" OR "bidirectional encoder representations from transformers" OR "AI language model") AND (medical practice OR healthcare OR clinical practice OR medicine OR medical treatment OR patient)

**Section 2. Excluded studies after full-text screening.**

| Title | Authors, publishing year | Journal | Included after initial screening | Excluded after full-text screening | Exclusion reason LH | Exclusion reason CR | Exclusion reason FB | Exclusion reason Table 1 |
|---|---|---|---|---|---|---|---|---|
| 1. Evaluating Risk Progression in Mental Health Chatbots Using Escalating Prompts | Heston, 2023 | medRxiv [Preprint] | LH, CR | LH, CR | aimed at detecting and addressing hazardous psychological states | restricted to psychology, psychosocial support, and behavior assessment | - | wrong discipline |
| 2. Chat GPT in Tailoring Individualized Lifestyle-Modification Programs in Metabolic Syndrome: Potentials and Difficulties? | Ismail, 2023 | Ann Biomed Eng | LH, CR | LH, CR | not original research | not original research | - | wrong study design |
| 3. Artificial Intelligence to Improve Patient Understanding of Radiology Report | Amin et al., 2023 | Yale J Biol Med | LH, CR | LH, CR | narrative review | not original research | - | wrong study design |
| 4. Enhancing Awareness and Self-diagnosis of Obstructive Sleep Apnea Using AI-Powered Chatbots: The Role of ChatGPT in Revolutionizing Healthcare | Bilal et al, 2023 | Ann Biomed Eng | LH, CR | LH, CR | not original research | not original research | - | wrong study design |
| 5. Deep learning for Arabic healthcare: MedicalBot | Abdelhay et al., 2023 | Soc Netw Anal Min | LH, CR | LH, CR | dataset and model development | technical development | - | technological development/perfor mance evaluation |
| 6. Examining Real-World Medication Consultations and Drug-Herb Interactions: ChatGPT Performance Evaluation | Hsu et al., 2023 | JMIR Med Educ | LH, CR | LH, CR | focus on pharmacological advice on drug interactions | investigating drug interactions | - | wrong discipline |
| 7. A Large Language Model Screening Tool to Target Patients for Best Practice Alerts: Development and Validation | Savage et al., 2023 | JMIR Med Inform | LH, CR | LH, CR | not directed to patients | wrong population | - | wrong population |
| 8. Mind + Machine: ChatGPT as a Basic Clinical Decisions Support Tool | Ayoub et al., 2023 | Cureus | LH, CR | LH, CR | not directed to patients | wrong population | - | wrong population |

| | | | | | | | | |
|---|---|---|---|---|---|---|---|---|
| 9. Personalized Impression Generation for PET Reports Using Large Language Models | Tie et al., 2024 | J Med Imaging Health Inform | LH, CR | LH, CR | not directed to patients | wrong population | - | wrong population |
| 10. Chatbots Vs. Human Experts: Evaluating Diagnostic Performance of Chatbots in Uveitis and the Perspectives on AI Adoption in Ophthalmology | Rojas-Carabali et al., 2023 | Ocul Immunol Inflamm | LH, CR | LH, CR | not directed to patients | wrong population | - | wrong population |
| 11. PERSONALIZING HEPATOLOGY: THE ROLE OF CHATBOTS IN TAILORED TREATMENT PLANS IN THE FIELD OF HEPATOLOGY | Rammohan et al., 2023 | Hepatology | LH, CR | LH, CR | abstract not providing sufficient information | insufficient data for thematic synthesis | - | insufficient data for thematic synthesis |
| 12. Artificial Intelligence for Kidney Stone Spectra Analysis: Using Artificial Intelligence Algorithms for Quality Assurance in the Clinical Laboratory | Day et al., 2023 | MCP: Digital Health | LH, CR | LH, CR | not directed to patients | wrong population | - | wrong population |
| 13. Physician and Patient Assessment of Extended Language Model Answers to Rheumatology Patient Inquiries: Doctor versus AI. Comment on the article by Ye et al | Daungsupawong et al., 2023 | Arthritis Rheumatol | LH | LH, CR | commentary | wrong study design | - | wrong study design |
| 14. A Hybrid Model for Depression Detection With Transformer and Bi-directional Long Short-Term Memory | Zhang et al., 2023 | IEEE BIBM 2023 | CR | LH, CR, FB | restricted to psychology, psychosocial support, and behavior assessment | wrong population | wrong discipline | wrong discipline |
| 15. Accurate Detection of Dementia from Speech Transcripts Using RoBERTa Model | Matošević et al., 2022 | MIPRO 2022 | CR | LH, CR, FB | restricted to psychology, psychosocial support, and behavior assessment | wrong population | wrong discipline | wrong discipline |
| 16. Detecting Reddit Users with Depression Using a | Chen et al., 2023 | arXiv [Preprint] | CR | LH, CR, FB | restricted to psychology, | wrong population | wrong discipline | wrong discipline |

| | | | | | | | | |
|---|---|---|---|---|---|---|---|---|
| Hybrid Neural Network SBERT-CNN | | | | | psychosocial support, and behavior assessment | | | |
| 17. BERT Learns From Electroencephalograms About Parkinson's Disease: Transformer-Based Models for Aid Diagnosis | Nogales et al., 2023 | IEEE Access | CR | LH, CR | not direct to patients | wrong population | - | wrong population |
| 18. The use of large language models in medicine: proceeding with caution | Deng et al., 2023 | Curr Med Res Opin | CR | LH, CR | commentary | wrong study design | - | wrong study design |
| 19. Identifying Common Topics in Patient Portal Messages with Unsupervised Natural Language Processing | Chang et al., 2023 | Int J Radiat. Oncol Biol Phys | LH | LH, CR | not direct to patients | wrong population | - | wrong population |
| 20. Democratizing access to research for patients with pancreatic cancer across a diverse health system through natural language processing of radiology reports | King et al., 2023 | J Clin Oncol | LH | LH, CR, FB | technical development | wrong population | technological development/perform ance evaluation | technological development/perfor mance evaluation |
| 21. Comparison of BERT implementations for natural language processing of narrative medical documents | Turchin et al., 2023 | Inform Med Unlocked | LH | LH, CR, FB | technical development | wrong population | technological development/perform ance evaluation | technological development/perfor mance evaluation |
| 22. Ensembles of BERT for Depression Classification | Senn et al., 2022 | Annu Int Conf IEEE Eng Med Biol Soc | LH, CR | LH, CR, FB | restricted to psychology, psychosocial support, and behavior assessment | wrong population | wrong discipline | wrong discipline |
| 23. One LLM is not Enough: Harnessing the Power of Ensemble Learning for Medical Question Answering | Yang et al., 2023 | medRxiv [Preprint] | LH, CR | LH, CR, FB | technical development | wrong population | technological development/perform ance evaluation | technological development/perfor mance evaluation |
| 24. FlauBERT vs. CamemBERT: Understanding patient's | Blanc et al., 2022 | Artif Intell Med | LH | LH, CR, FB | technical development | wrong population | technological development/perform ance evaluation | technological development/perfor mance evaluation |

| | | | | | | | | |
|---|---|---|---|---|---|---|---|---|
| answers by a French medical chatbot | | | | | | | | |
| 25. ChatGPT-4 and the Global Burden of Disease Study: Advancing Personalized Healthcare Through Artificial Intelligence in Clinical and Translational Medicine | Temsah et al., 2023 | Cureus | LH | LH, CR | editorial | not original research | - | wrong study design |
| 26. Can we use ChatGPT for Mental Health and Substance Use Education? Examining Its Quality and Potential Harms | Spallek et al., 2023 | JMIR Med Educ | CR | LH, CR | restricted to psychology, psychosocial support, and behavior assessment | focus on mental health and substance use | - | wrong discipline |
| 27. Advancing Mental Health Diagnostics: GPT-Based Method for Depression Detection | Danner et al., 2023 | SICE2023 | CR | LH, CR | restricted to psychology, psychosocial support, and behavior assessment | focus on psychology | - | wrong discipline |
| 28. Large language models in bariatric surgery patient support: A transformative approach to patient education and engagement | Samaan et al., 2023 | Clin Obes | LH, CR | LH, CR | correspondence | not original research | - | wrong study design |
| 29. Application of Large Language Models such as ChatGPT to Support Nutritional Recommendations for Dialysis Patients | Wang et al., 2023 | Kidney Week 2023 | LH, CR | LH, CR | abstract not providing sufficient information | insufficient data for thematic synthesis | - | insufficient data for thematic synthesis |
| 30. Enabling the Informed Patient Paradigm with Secure and Personalized Medical Question Answering | Oduro-Afriyie et al., 2023 | ACM-BCB 2023 | LH, CR | LH, CR, FB | technical development | wrong population | technological development/performance evaluation | technological development/performance evaluation |
| 31. Exploring the Potential of Chat GPT in Personalized Obesity Treatment | Arslan, 2023 | Ann Biomed Eng | LH, CR | LH, CR | narrative review | not original research | - | wrong study design |
| 32. O16 Can sexual health clinicians be replaced by robots? Utility of artificial | Taylor et al., 2023 | BASHH 2023 | LH, CR | LH, CR | abstract not providing sufficient information | insufficient data for thematic synthesis | - | insufficient data for thematic synthesis |

| | | | | | | | | |
|---|---|---|---|---|---|---|---|---|
| intelligence platforms to give sexual health advice | | | | | | | | |
| 33. O-089 Using ChatGPT to answer patient questions about fertility: the quality of information generated by a deep learning language model | Beilby et al., 2023 | 39th Hybrid Annual Meeting of the ESHRE | LH, CR | LH, CR | abstract not providing sufficient information | insufficient data for thematic synthesis | - | insufficient data for thematic synthesis |
| 34. Enhancing Kidney Transplant Care through the Integration of Chatbot | Valencia et al., 2023 | Healthcare | LH, CR | LH, CR | narrative review | not original research | - | wrong study design |
| 35. Assessing the accuracy and completeness of artificial intelligence language models in providing information on methotrexate use | Coskun et al., 2023 | Rheumatol Int | LH, CR | LH, CR | restricted to pharmaceutical treatment | focus on MTX | - | wrong discipline |

### Section 3A. Presence of codes for included studies number 1-45.

| First order codes | Second order codes | Third order codes | 1. Samaan et al. | 2. Eromosele et al. | 3. Johri et al. | 4. Braga et al. | 5. King et al. | 6. Huang et al. | 7. Hanna et al. | 8. Liu et al. | 9. Samaan et al. | 10. Patnaik et al. | 11. Ali et al. | 12. Suresh et al. | 13. Yeo et al. | 14. Knebel et al. | 15. Zhu et al. | 16. Lahat et al. | 17. Bernstein et al. | 18. Rogasch et al. | 19. Campbell et al. | 20. Currie et al. | 21. Draschl et al. | 22. Alessandri-Bonetti et al. | 23. Capelleras et al. | 24. Coskun et al. | 25. Durairaj et al. | 26. Kianian et al. | 27. Seth et al. | 28. Inojosa et al. | 29. Lyons et al. | 30. Babayiğit et al. | 31. Mondal et al. | 32. Kim et al. | 33. Song et al. | 34. Bitar et al. | 35. Zalzal et al. | 36. Chervenak et al. | 37. Bushuven et al. | 38. Jeblick et al. | 39. Samaan et al. | 40. Zhou et al. | 41. Oniani et al. | 42. Hernandez et al. | 43. Kuşçu et al. | 44. Biswas et al. | 45. Chiesa-Estomba et al. |
|---|---|---|---|---|---|---|---|---|---|---|---|---|---|---|---|---|---|---|---|---|---|---|---|---|---|---|---|---|---|---|---|---|---|---|---|---|---|---|---|---|---|---|---|---|---|---|---|
| Applications | | | | | | | | | | | | | | | | | | | | | | | | | | | | | | | | | | | | | | | | | | | | | | | |
| Language [other than English] | | | | | | | | | | | 1 | | | | 1 | | | | | | | | | | | | | | | | | | | | | | | | | | | | | | | | |
| | Arab | | | | | | | | | | 1 | | | | | | | | | | | | | | | | | | | | | | | | | | | | | | | | | | | | |
| | Korean | | | | | | | | | | | | | | 1 | | | | | | | | | | | | | | | | | | | | | | | | | | | | | | | | |
| | Mandarin | | | | | | | | | | | | | | 1 | | | | | | | | | | | | | | | | | | | | | | | | | | | | | | | | |
| | Spanish | | | | | | | | | | | | | | 1 | | | | | | | | | | | | | | | | | | | | | | | | | | | | | | | | |
| Discipline | | | 1 | 1 | 1 | 1 | 1 | 1 | 1 | 1 | 1 | 1 | 1 | 1 | 1 | 1 | 1 | 1 | 1 | 1 | 1 | 1 | 1 | 1 | 1 | 1 | 1 | 1 | 1 | 1 | 1 | 1 | 1 | 1 | 1 | 1 | 1 | 1 | 1 | 1 | 1 | 1 | 1 | 1 | 1 | 1 | 1 |
| | Anesthesiology | | | | | | | | | | | 1 | | | | | | | | | | | | | | | | | | | | | | | | | | | | | | | | | | | |
| | Cardiology | | | 1 | | | 1 | | | | | | | | | | | | | | | | | | | | | | | | | | | | | | | | | | | | | | | | |
| | Dentistry | | | | | | | | | | | | | | | | | | | | | | | | | | | | | | | 1 | | | | | | | | | | | | | | | |
| | Dermatology | | | | 1 | | | | | | | | | | | | | | | | | | | | | | | | | | | | | | | | | | | | | | | | | | |
| | Emergency Medicine | | | | | | | | | | | | | | | | | | | | | | | | | | | | | | | | | | | | | | 1 | | | | | | | | |
| | Endocrinology | | | | | | | | | | | | | | | | | | | | | | | | | | | | | | | | 1 | | | | | | | | | | | 1 | | | |
| | Gastroenterology | | 1 | | | | | | | | 1 | | 1 | | 1 | | | 1 | | | | | | | | | | | | | | | | | | | | | | | | 1 | 1 | | | | |
| | General Surgery | | | | | | | | | | | | | | | | | | | | | | | | | | | | | | | | | | | | | | | | 1 | | | | | | |
| | Gynecology | | | | | | | | | | | | | | | | | | | | | | | | | | | | | | | | | | | 1 | | | | | | | | | | | |
| | Hand Surgery | | | | | | | | | | | | | | | | | | | | | | | | | | | 1 | | | | | | | | | | | | | | | | | | | |
| | Head and Neck Surgery/Otolaryngology | | | | | | | | | | | | | 1 | | | | | | | 1 | | | | | | | | | | | | | | | | 1 | | | | | | | | 1 | | 1 |
| | Infectious Disease | | | | | | | | 1 | | | | | | | | | | | | | | | | | | | | | | | | | | | | | | | | | | | | | | |
| | Nephrology | | | | | | | | | | | | | | | | | | | | | | | | | | | | | | | | | | | | | | | | | | | | | | |
| | Neurology | | | | | | | 1 | | | | | | | | | | | | | | | | | | | | | 1 | | | | 1 | | | | | | | | | | | | | | |
| | Neurosurgery | | | | | | | | | | | | | | | | | | | | | | | | | | | | | | | | | | | | | | | | | | | | | | |
| | Nuclear Medicine | | | | | | | | | | | | | | | | | | 1 | | 1 | | | | | | | | | | | | | | | | | | | | | | | | | | |
| | Oncology | | | | | | | | | | | | | | | | | | | | | | | | | | | | | | | | | | | | | | | | | | | | | | |
| | Ophthalmology | | | | | | | | | | | | | | 1 | | | 1 | | | | | | | | | 1 | | | 1 | | | | | | | | | | | | | | | 1 | |

| | | | | | | | | | | | | | | | | | | | | | | | | | | | | | | | | | | | | | | | | | | | | | | |
|---|---|---|---|---|---|---|---|---|---|---|---|---|---|---|---|---|---|---|---|---|---|---|---|---|---|---|---|---|---|---|---|---|---|---|---|---|---|---|---|---|---|---|---|---|---|---|---|
| | Orthopedics | | | | | | | | | | | | | | | | | | | | | | | | 1 | | | | | | | | | | | | | | | | | | | | | | |
| | Plastic Surgery | | | | | | | | | | | | | | | | | | | | | | | | | 1 | 1 | | 1 | | | | | | | | | | | | | | | | | | |
| | Primary Care/General | | | | | | | | | 1 | | | | | | | | | | | | | | | | | | | | | | | | | | | | | | | | | | | | | |
| | Pulmonology | | | | | | | | | | | | | | | | | | | | | | | | | | | | | | | | | | | | | | | | | | | | | | |
| | Radiation Oncology | | | | | | | | | | | | | | | | | | | | | | | | | | | | | | | | | | | | | | | | | | | | | | |
| | Radiology | | | | | | | | | | | | | | | | | | | | | | | | | | | | | | | | | | | | | | 1 | | | | | | | | |
| | Reproductive Medicine | | | | | | | | | | | | | | | | | | | | | | | | | | | | | | | | | | | | 1 | | | | | | | | | | |
| | Rheumatology | | | | | | | | | | | | | | | | | | | | | | | | | | | | | | | | | | | | | | | | | | | | | | |
| | Thoracic Surgery | | | | | | | | | | | | | | | | | | | | | | | | | | | | | | | | | | | | | | | | | | | | | | |
| | Urology | | | | | 1 | | | | | | | | | | | 1 | | | | | | | | | | | 1 | | | | | | | | | 1 | | | | | | | | | | |
| | Vascular Surgery | | | | | | | | | | | | | | | | | | | | | | | | | | | | | | | | | | | | | | | | | | | | | | |
| Medical question answering/chatbot | | | 1 | 1 | 1 | 1 | 1 | 1 | 1 | 1 | 1 | 1 | 1 | 1 | 1 | 1 | 1 | 1 | 1 | 1 | 1 | 1 | 1 | 1 | 1 | 1 | 1 | 1 | 1 | 1 | 1 | 1 | 1 | 1 | 1 | 1 | 1 | 1 | 1 | 1 | 1 | 1 | | 1 | 1 | 1 | 1 |
| Generation of patient information | | | | | | | | | 1 | | | | | | | | | | | | | | 1 | | | | | | 1 | 1 | | | | | | | 1 | | 1 | | 1 | | | 1 | | | |
| | Clinical documentation | | | | | | | | 1 | | | | | | | | | | | | | | 1 | | | | | | | | | | | | | | | | | | | | | | | | |
| | | Informed consent | | | | | | | | | | | | | | | | | | | | 1 | | | | | | | | | | | | | | | | | | | | | | | | | |
| | | Discharge instructions | | | | | | | 1 | | | | | | | | | | | | | | | | | | | | | | | | | | | | | | | | | | | | | | |
| | Medical text summarization/translation | | | | | | | | | | | | | | | | | | | | | | | | | | | 1 | 1 | | | | | | | | 1 | | 1 | | 1 | | | 1 | | | |
| | | Patient education materials | | | | | | | | | | | | | | | | | | | | | | | | | | 1 | | | | | | | | | | | | | | | 1 | | | | |
| | | Simplified radiology reports | | | | | | | | | | | | | | | | | | | | | | | | | | | | | | | | | | | | | | | 1 | | | | | | |
| | | Patient-friendly medical responses/summaries | | | | | | | | | | | | | | | | | | | | | | | | | | | 1 | | | | | | | 1 | | 1 | | | | | | | | | |
| Disease management and support | | | 1 | 1 | 1 | 1 | 1 | | 1 | 1 | 1 | 1 | 1 | 1 | 1 | 1 | 1 | 1 | 1 | 1 | 1 | 1 | 1 | 1 | 1 | 1 | 1 | 1 | 1 | 1 | 1 | 1 | 1 | 1 | 1 | 1 | 1 | 1 | 1 | 1 | 1 | 1 | 1 | 1 | 1 | 1 | 1 |
| | Prevention | | 1 | 1 | | | 1 | | | 1 | 1 | | | 1 | 1 | | 1 | | | | | | 1 | | | | | | | | | 1 | | | | 1 | | | | 1 | 1 | 1 | 1 | | | | |

| | | | | | | | | | | | | | | | | | | | | | | | | | | | | | | | | | | | | | | | | | | | | | | |
|---|---|---|---|---|---|---|---|---|---|---|---|---|---|---|---|---|---|---|---|---|---|---|---|---|---|---|---|---|---|---|---|---|---|---|---|---|---|---|---|---|---|---|---|---|---|---|---|
| | Prehospital management /triage | | | | | | | | | | | | | | | 1 | | | | | | | | | | | | | | | 1 | | 1 | | | | | | | | | | | | | | |
| | Diagnosis | | | 1 | 1 | 1 | | | 1 | 1 | 1 | | | 1 | 1 | 1 | 1 | 1 | 1 | 1 | 1 | | 1 | | | 1 | | | 1 | 1 | 1 | | 1 | 1 | 1 | 1 | 1 | | 1 | 1 | | 1 | 1 | | | | |
| | Treatment | | 1 | 1 | | 1 | | | 1 | 1 | 1 | 1 | 1 | 1 | 1 | | 1 | 1 | 1 | 1 | 1 | 1 | 1 | 1 | 1 | 1 | 1 | 1 | 1 | 1 | | 1 | | 1 | 1 | 1 | | 1 | 1 | 1 | | 1 | 1 | 1 | 1 | 1 | 1 |
| | Prognosis | | | 1 | | | | | | | | 1 | | | | | | | | | | | | | | 1 | | | | 1 | | | | | | | | 1 | | 1 | 1 | | 1 | | | | 1 |
| **Limitations** | | | | | | | | | | | | | | | | | | | | | | | | | | | | | | | | | | | | | | | | | | | | | | | |
| Design | | | 1 | 1 | 1 | 1 | 1 | 1 | | 1 | 1 | | 1 | 1 | 1 | 1 | 1 | 1 | 1 | 1 | 1 | 1 | | 1 | | 1 | 1 | | 1 | 1 | 1 | | 1 | 1 | 1 | | 1 | 1 | 1 | 1 | 1 | 1 | 1 | | 1 | 1 | 1 |
| | Not optimized for the medical domain | | | 1 | | 1 | | | | 1 | | | 1 | 1 | | 1 | 1 | 1 | | | | | | | | 1 | | | 1 | 1 | | | | 1 | 1 | | 1 | 1 | 1 | 1 | 1 | | 1 | 1 | | | 1 | 1 |
| | | Implicit knowledge/lack of clinical context | | | | 1 | | | | | | | | | | | | 1 | | | | | | | | 1 | | | | | | | | | | | | | | | | | | | | | 1 |
| | | Limited clinical reasoning | | | | | | | | | | | | | | | | | | | | | | | | | | | | | | | | | | | 1 | | | | | | | | | | |
| | | Limited in processing/producing medical images | | | | | | | | | | | | | | 1 | | | | | | | | | | | | | | | | | | | 1 | | | | | | | | | | | | |
| | | Misunderstanding of medical information/terms | | | | 1 | | | | | | | | | | | 1 | 1 | | | | | | | | | | | | | | | | | | | | | 1 | | | 1 | | | 1 | |
| | Data | | 1 | 1 | | | 1 | 1 | | 1 | 1 | | 1 | | 1 | 1 | 1 | 1 | 1 | 1 | | 1 | | 1 | | 1 | 1 | | 1 | 1 | 1 | | 1 | 1 | 1 | | 1 | 1 | 1 | 1 | 1 | | 1 | | 1 | 1 | |
| | | Restricted access to internet data | | | | | | | | | | | | | | | 1 | 1 | | 1 | | 1 | | | | | | | | | | | | 1 | 1 | | 1 | 1 | | 1 | 1 | | | | 1 | | |
| | | Undisclosed origin of training data | 1 | 1 | | | 1 | 1 | | | 1 | | 1 | | 1 | 1 | | | 1 | | | | | | | 1 | 1 | | | 1 | 1 | | 1 | 1 | 1 | | 1 | 1 | 1 | 1 | 1 | | | | 1 | 1 | |
| | | Limited in reference provision/evaluation/validation | | | | | | | | | | | | | | | | | | | | | | 1 | | | | | | 1 | | | | | 1 | 1 | | 1 | 1 | | | | | | | | 1 | |
| | | Stores/processes sensitive health information | | | | | | | | 1 | | | 1 | | | | | | | | | | | | | 1 | | | | | | | | | 1 | | | | | | | | 1 | | | | |
| | Black box | | | | 1 | | | | | | | | | | 1 | | | | | | | | | | | | | | | | | | | | 1 | | | 1 | | | | | | | | 1 | |
| | Limited engagement/dialogue capabilities | | | | 1 | 1 | | | | 1 | | | | | | 1 | 1 | | | | | | | | | | | | | 1 | | | | | | | 1 | | | | | | | | | | 1 |

| | | | | | | | | | | | | | | | | | | | | | | | | | | | | | | | | | | | | | | | | | | | | | | | |
|---|---|---|---|---|---|---|---|---|---|---|---|---|---|---|---|---|---|---|---|---|---|---|---|---|---|---|---|---|---|---|---|---|---|---|---|---|---|---|---|---|---|---|---|---|---|---|---|
| | Incapable of self-validation/correction | | | | | | | | | | | | | | | | | | | | | | | | | | | | | | | | | | | | | | | | | 1 | | | | | |
| | Accessibility | | | | 1 | | | | | | | | 1 | | 1 | | | | | | 1 | 1 | | | | | | | | | | | | 1 | | | 1 | | | | 1 | | | 1 | 1 | |
| | | Limited number of prompts | | | | | | | | | | | 1 | | | | | | | | | | | | | | | | | | | | | | | | | | | | | | | 1 | | |
| | | Not freely accessible | | | | | | | | | | | 1 | | | | | | | | 1 | 1 | | | | | | | | | | | | | | | | | | | 1 | | | 1 | | |
| | | Not open access | | | 1 | | | | | | | | | | 1 | | | | | | | | | | | | | | | | | | | 1 | | | 1 | | | | | | | | 1 | |
| Output | | | 1 | 1 | 1 | 1 | 1 | 1 | 1 | 1 | 1 | 1 | 1 | 1 | 1 | 1 | 1 | 1 | 1 | 1 | 1 | 1 | 1 | 1 | 1 | 1 | | 1 | 1 | 1 | 1 | 1 | 1 | 1 | 1 | | 1 | 1 | 1 | 1 | 1 | 1 | 1 | | 1 | 1 | 1 |
| | Non-reproducible | | | | | 1 | 1 | | | | | | 1 | | | | 1 | 1 | | 1 | | 1 | | 1 | | 1 | | | 1 | | | | | 1 | 1 | | 1 | 1 | 1 | 1 | 1 | 1 | | | 1 | 1 | |
| | | Non-deterministic | | | | 1 | 1 | | | | | | 1 | | | | 1 | 1 | | 1 | | 1 | | | | 1 | | | | | | | | | | | | 1 | 1 | 1 | 1 | | | 1 | | |
| | | Non-referenceable | | | | | | | | | | | | | | | | | | | | | | 1 | | | | | 1 | | | | | 1 | 1 | | 1 | 1 | | | | | | | 1 | |
| | Non-comprehensive | | 1 | 1 | | 1 | 1 | 1 | | 1 | 1 | 1 | 1 | 1 | 1 | 1 | 1 | 1 | 1 | 1 | 1 | 1 | 1 | | | 1 | | 1 | 1 | 1 | 1 | 1 | 1 | 1 | 1 | | 1 | 1 | 1 | 1 | 1 | 1 | 1 | | 1 | 1 | |
| | | Generic/non-personalized | | | | 1 | | 1 | | 1 | | | 1 | | | 1 | 1 | | | 1 | 1 | 1 | | | | | | | 1 | | 1 | | | | | | 1 | 1 | | 1 | | 1 | | | | 1 | |
| | | Incomplete | 1 | 1 | | 1 | 1 | 1 | | 1 | 1 | | 1 | 1 | 1 | 1 | 1 | 1 | | 1 | 1 | 1 | 1 | | | 1 | | | 1 | 1 | 1 | 1 | | | 1 | | 1 | 1 | 1 | 1 | 1 | 1 | 1 | | 1 | 1 | |
| | | Non-standard of care | | | | 1 | | | | | | | | | | | | | 1 | | | 1 | | | | 1 | | | 1 | 1 | | | | 1 | | | | 1 | 1 | | | | | | | 1 | |
| | | Outdated | 1 | | | | | | | 1 | 1 | | 1 | | | | 1 | | | 1 | | 1 | 1 | | | | | | 1 | | | | | 1 | | | | | | | | | | | | | |
| | | Oversimplification | | | | | | | | | | | | | | | 1 | | | | | | | | | 1 | | | 1 | | | | | 1 | | | | | | 1 | | | | | | | |
| | | Superfluous | | | | 1 | | | | 1 | | | 1 | | | | | | | | | | | | | 1 | | | | | | | | | | | | | | | | 1 | 1 | | | | |
| | | Overcautious | | | | 1 | | | | 1 | | | | | | 1 | | | | | | | | | | | | | | | 1 | | | | | | | | | | | | | | | | |
| | | Overempathic | | | | | | | | 1 | | | | | | | | | | | | | | | | | | | | | | | | | | | | | | | | | | | | | |
| | | High complexity/reading level | | | | | | | | 1 | | 1 | | | | | | | | | 1 | | | | | | | 1 | | 1 | 1 | | 1 | | 1 | | 1 | | | | | | | | | | |
| | Incorrect | | 1 | 1 | 1 | 1 | 1 | 1 | 1 | 1 | 1 | 1 | 1 | 1 | 1 | 1 | 1 | 1 | 1 | 1 | 1 | 1 | 1 | | | 1 | | | 1 | 1 | 1 | 1 | | 1 | 1 | | 1 | 1 | 1 | 1 | 1 | 1 | 1 | | 1 | 1 | 1 |
| | | Extrapolation | | | | | | | | | | | | | | | | | | | | 1 | | | | | | | | | | | | | | | | | | 1 | | | | | | 1 | |
| | | Delirium | | 1 | | 1 | | 1 | | 1 | | | | | | 1 | | | | | | 1 | | | | | | | | 1 | | | | | | | | | 1 | 1 | | 1 | | | | 1 | |
| | | Delusion | | | | 1 | | 1 | | | | | | | | | | | | | | 1 | | | | | | | | 1 | | | | | | | | | | 1 | | | | | | 1 | |
| | | Hallucination | 1 | | | 1 | | | | | 1 | 1 | | 1 | 1 | 1 | 1 | | 1 | 1 | 1 | 1 | 1 | | | | | | 1 | 1 | 1 | | | | | | | 1 | 1 | 1 | 1 | | | | | 1 | |
| | | Illusion | | | | 1 | | | | | | | | | 1 | | 1 | | | | | 1 | | | | | | | | | | | | | | | | 1 | | 1 | | | | | | | |
| | | Confabulation | 1 | | | 1 | | | | | | | | | 1 | 1 | 1 | | 1 | | | | | | | 1 | | | | | | | | | | | | | | 1 | | | 1 | | | 1 | |
| | Unsafe | | | | | 1 | | 1 | | | | | | 1 | | 1 | | 1 | 1 | | 1 | 1 | 1 | | | 1 | | | | 1 | 1 | | | | | | | 1 | 1 | 1 | 1 | | 1 | | 1 | 1 | |
| | | Misleading | | | | 1 | | 1 | | | | | | 1 | | | | | | | | 1 | 1 | | | 1 | | | | 1 | 1 | | | | | | | 1 | 1 | 1 | 1 | | 1 | | 1 | 1 | |
| | | Harmful | | | | 1 | | 1 | | | | | | | | 1 | | 1 | 1 | | 1 | 1 | | | | | | | | 1 | 1 | | | | | | | | 1 | 1 | 1 | | | | | | |

| | | | | | | | | | | | | | | | | | | | | | | | | | | | | | | | | | | | | | | | | | | | | | | | |
|---|---|---|---|---|---|---|---|---|---|---|---|---|---|---|---|---|---|---|---|---|---|---|---|---|---|---|---|---|---|---|---|---|---|---|---|---|---|---|---|---|---|---|---|---|---|---|---|
| | Prompt-/input dependent | | | 1 | 1 | 1 | | | | | | | 1 | | | | 1 | | | 1 | | | 1 | | | 1 | | | | | 1 | 1 | | | | | 1 | | | | | | | | | | |
| | | Evidence | | | | | | | | | | | | | | | | | | | | | | | | | | | | | | 1 | | | | | | | | | | | | | | | |
| | | Complexity | | | | 1 | | | | | | | 1 | | | | | | | | | | 1 | | | 1 | | | | | | | | | | | | | | | | | | | | | |
| | | Specificity | | | 1 | | | | | | | | | | | | 1 | | | 1 | | | | | | | | | | | | | | | | | 1 | | | | | | | | | | |
| | | Quantity | | 1 | | | | | | | | | | | | | | | | | | | | | | | | | | | | 1 | | | | | | | | | | | | | | | |
| | | Conversation type | | | 1 | | | | | | | | | | | | | | | | | | | | | | | | | | 1 | | | | | | | | | | | | | | | | |
| | Environment-dependent | | | | | | | | | | 1 | | | 1 | | | | | | | | | | | | 1 | | | 1 | 1 | 1 | | | 1 | | | | | 1 | | 1 | | | | | | |
| | | Target group | | | | | | | | | | | | | | | | | | | | | | | | 1 | | | 1 | | 1 | | | 1 | | | | | | | | | | | | | |
| | | Provider/organization | | | | | | | | | 1 | | | | | | | | | | | | | | | | | | | 1 | | | | | | | | | | | 1 | | | | | | |
| | | Local/national medical resources | | | | | | | | | | | | 1 | | | | | | | | | | | | | | | | 1 | | | | | | | | | 1 | | 1 | | | | | | |
| | Bias | | | 1 | | | | | | | | 1 | | 1 | | 1 | | | | | | | | | | | | | | | | | | | | | | | | | | | | | | | 1 |
| | | Language | | | | | | | | | | 1 | | | | 1 | | | | | | | | | | | | | | | | | | | | | | | | | | | | | | | |
| | | Insurance status | | | | | | | | | | | | | | | | | | | | | | | | | | | | | | | | | | | | | | | | | | | | | |
| | | Underserved racial groups | | 1 | | | | | | | | | | | | | | | | | | | | | | | | | | | | | | | | | | | | | | | | | | | |
| | | Underrepresented procedures | | | | | | | | | | | | 1 | | | | | | | | | | | | | | | | | | | | | | | | | | | | | | | | | |

### Section 3B. Presence of codes for included studies number 46-89.

| First order codes | Second order codes | Third order codes | 46. Decker et al. | 47. Kaarre et al. | 48. Ferreira et al. | 49. Truhn et al. | 50. Hurley et al. | 51. Cankurtaran et al. | 52. Birkun et al. | 53. Pushpanathan et al. | 54. Shao et al. | 55. Vaira et al. | 56. Chen et al. | 57. Bellinger et al. | 58. Nielsen et al. | 59. Sezgin et al. | 60. Floyd et al. | 61. Uz et al. | 62. Athavale et al. | 63. Li et al. | 64. Seth et al. | 65. Kuckelman et al. | 66. Lockie et al. | 67. Harver et al. | 68. Li et al. | 69. Scheschenja et al. | 70. Gordon et al. | 71. Stroop et al. | 72. Coraci et al. | 73. Ye et al. | 74. Mohammad-Rahimi et al. | 75. Scquizzato et al. | 76. Hermann et al. | 77. Kerbage et al. | 78. Shiraishi et al. | 79. Barclay et al. | 80. Qarajeh et al. | 81. Chowdhury et al. | 82. Singer et al. | 83. Xie et al. | 84. Nastasi et al. | 85. Sulejmani et al. | 86. Biswas et al. | 87. Panagoulias et al. | 88. Chandra et al. | 89. Hung et al. | Total codes |
|---|---|---|---|---|---|---|---|---|---|---|---|---|---|---|---|---|---|---|---|---|---|---|---|---|---|---|---|---|---|---|---|---|---|---|---|---|---|---|---|---|---|---|---|---|---|---|---|
| **Applications** | | | | | | | | | | | | | | | | | | | | | | | | | | | | | | | | | | | | | | | | | | | | | | | |
| Language [other than English] | | | | | | | | | | | 1 | | | | | | | | | | | | | | | | | | | | | | | | | | | | | | | | 1 | | | | 4 |
| | Arab | | | | | | | | | | | | | | | | | | | | | | | | | | | | | | | | | | | | | | | | | | 1 | | | | 2 |
| | Korean | | | | | | | | | | | | | | | | | | | | | | | | | | | | | | | | | | | | | | | | | | | | | | 1 |
| | Mandarin | | | | | | | | | | 1 | | | | | | | | | | | | | | | | | | | | | | | | | | | | | | | | | | | | 2 |
| | Spanish | | | | | | | | | | | | | | | | | | | | | | | | | | | | | | | | | | | | | | | | | | | | | | 1 |
| Discipline | | | 1 | 1 | 1 | 1 | 1 | 1 | 1 | 1 | 1 | 1 | 1 | 1 | 1 | 1 | 1 | 1 | 1 | 1 | 1 | 1 | 1 | 1 | 1 | 1 | 1 | 1 | 1 | 1 | 1 | 1 | 1 | 1 | 1 | 1 | 1 | 1 | 1 | 1 | 1 | 1 | 1 | 1 | 1 | 1 | 1 | 89 |
| | Anesthesiology | | | | | | | | | | | | | | | | | | | | | | | | | | | | | | | | | | | | | | | | | | | | | | 1 |
| | Cardiology | | | | | | | | | | | | | | | | | | | | | | | | | | | | | | | | | | | | | | | | | | | | | | 3 |
| | Dentistry | | | | | | | | | | | | | | | | | | | | | | | | | | | | | | 1 | | | | | | | | | | | | | | | | 2 |
| | Dermatology | | | | 1 | | | | | | | | | | | | | | | | | | | | | | | | | | | | | | | | | | | | | 1 | | | 1 | | 4 |
| | Emergency Medicine | | | | | | | | 1 | | | | | | | | | | | | | | | | | | | | | | | 1 | | | | | | | | | | | | | | | 3 |
| | Endocrinology | | | | | | | | | | | | | | | | | | | | | | | | | | | | | | | | | | | | | | | | | | | | | | 2 |
| | Gastroenterology | | | | | | | 1 | | | | | | | | | | | | | | | | | | | | | | | | | | 1 | | | | | | | | | | | | | 9 |
| | General Surgery | | 1 | | | | | | | | | | | | | | | | | | | | 1 | | | | | | | | | | | | | | | | | | | | | | | | 3 |
| | Gynecology | | | | | | | | | | | | | | | 1 | | | | | | | | | | | | | | | | | 1 | | | | | | | | | | | | | | 3 |
| | Hand Surgery | | | | | | | | | | | | | | | | | | | | | | | | | | | | | | | | | | | | | | | | | | | | | | 1 |
| | Head and Neck Surgery/Otolaryngology | | | | | | | | | | | 1 | | 1 | 1 | | | | | | | | | | | | | | | | | | | | | | | | | | | | | | | | 8 |
| | Infectious disease | | | | | | | | | | | | | | | | | | | | | | | | | | | | | | | | | | | | | | | | | | | | | | 1 |
| | Nephrology | | | | | | | | | | | | | | | | | | | | | | | | | | | | | | | | | | | | 1 | | | | | | | | | | 1 |
| | Neurology | | | | | | | | | | | | | | | | | | | | | | | | | | | | | | | | | | | | | | | | | | | | | | 3 |
| | Neurosurgery | | | | | | | | | | | | | | | | | | | | | | | | | | 1 | | | | | | | | | | | | | | | | | | | | 1 |
| | Nuclear Medicine | | | | | | | | | | | | | | | | | | | | | | | | | | | | | | | | | | | | | | | | | | | | | | 2 |
| | Oncology | | | | | | | | | | | | 1 | | | | | | | | | | | | | | | | | | | | | | | | | | | | | | | | | | 1 |
| | Ophthalmology | | | | | | | | | 1 | | | | | | | | | | | | | | | | | | | | | | | | | 1 | 1 | | 1 | 1 | | | | | | | | 10 |

| | | | | | | | | | | | | | | | | | | | | | | | | | | | | | | | | | | | | | | | | | | | | | | | |
|---|---|---|---|---|---|---|---|---|---|---|---|---|---|---|---|---|---|---|---|---|---|---|---|---|---|---|---|---|---|---|---|---|---|---|---|---|---|---|---|---|---|---|---|---|---|---|---|
| | Orthopedics | | | 1 | | 1 | 1 | | | | | | | | | | | | | | | | | | | | | | 1 | | | | | | | | | | | | | | | | | | 5 |
| | Plastic Surgery | | | | | | | | | | | | | | | | | | | | 1 | | | | | | | | | | | | | | | | | | | | 1 | | | | | | 1 | 6 |
| | Primary Care/General | | | | | | | | | | | | | | | | | | | 1 | | | | | | | | | | | | | | | | | | | | | | 1 | | 1 | | | | 4 |
| | Pulmonology | | | | | | | | | | | | | | | | | | | | | | | | 1 | | | | | | | | | | | | | | | | | | | | 1 | | | 2 |
| | Radiation Oncology | | | | | | | | | | | | | | | | 1 | | | | | | | | | | | | | | | | | | | | | | | | | | | | | | | 1 |
| | Radiology | | | | | 1 | | | | | | | | | | | | | | | | | 1 | | | 1 | 1 | 1 | | | | | | | | | | | | | | | | | | | | 6 |
| | Reproductive Medicine | | | | | | | | | | | | | | | 1 | | | | | | | | | | | | | | | | | | | | | | | | | | | | | | | | 2 |
| | Rheumatology | | | | | | | | | | | | | | | | | 1 | | | | | | | | | | | | | | 1 | | | | | | | | | | | | | | | | 2 |
| | Thoracic Surgery | | | | | | | | | | 1 | | | | | | | | | | | | | | | | | | | | | | | | | | | | | | | | | | | | | 1 |
| | Urology | | | | | | | | | | | | | | | | | | | | | | | | | | | | | | | | | | | | | | | | | | | | | | | 4 |
| | Vascular Surgery | | | | | | | | | | | | | | | | | | 1 | | | | | | | | | | | | | | | | | | | | | | | | | | | | | 1 |
| Medical question answering/chatbot | | | | 1 | 1 | | 1 | 1 | 1 | 1 | 1 | 1 | 1 | 1 | 1 | 1 | 1 | 1 | 1 | 1 | 1 | 1 | 1 | 1 | 1 | 1 | 1 | 1 | 1 | 1 | 1 | 1 | 1 | 1 | | 1 | 1 | 1 | 1 | 1 | 1 | 1 | 1 | 1 | 1 | | 84 |
| Generation of patient information | | | 1 | | | 1 | | | | | | | | | 1 | | | | | | | | | | | 1 | 1 | 1 | 1 | | | | | | | 1 | | | | | 1 | | | | | 1 | 1 | 19 |
| | Clinical documentation | | 1 | | | | | | | | | | | | | | | | | | | | | | | | | | 1 | | | | | | | 1 | | | | | 1 | | | | | | | 6 |
| | | Informed consent | 1 | | | | | | | | | | | | | | | | | | | | | | | | | | 1 | | | | | | | 1 | | | | | 1 | | | | | | | 5 |
| | | Discharge instructions | | | | | | | | | | | | | | | | | | | | | | | | | | | | | | | | | | | | | | | | | | | | | | 1 |
| | Medical text summarization/translation | | | | | | | | | | | | | | 1 | | | | | | | | | | | 1 | 1 | 1 | | | | | | | | | | | | | | | | | | 1 | 1 | 12 |
| | | Patient education materials | | | | | | | | | | | | | | | | | | | | | | | | | | 1 | | | | | | | | | | | | | | | | | | 1 | 1 | 5 |
| | | Simplified radiology reports | | | | | | | | | | | | | | | | | | | | | | | | 1 | | | | | | | | | | | | | | | | | | | | | | 2 |
| | | Patient-friendly medical responses/summaries | | | | | | | | | | | | | 1 | | | | | | | | | | | | 1 | | | | | | | | | | | | | | | | | | | | | 5 |
| Disease management and support | | | 1 | 1 | 1 | 1 | 1 | 1 | 1 | 1 | 1 | | | 1 | 1 | 1 | 1 | 1 | 1 | 1 | 1 | 1 | 1 | 1 | 1 | 1 | 1 | 1 | 1 | 1 | 1 | 1 | 1 | 1 | 1 | 1 | 1 | 1 | 1 | 1 | 1 | 1 | 1 | 1 | 1 | 1 | 86 |
| | Prevention | | | | | | | 1 | | | 1 | | | | | | | | | | | | | 1 | | | | | | | 1 | | 1 | | | | | | | | 1 | | 1 | | | | | 22 |
| | Prehospital management/triage | | | | | 1 | | | 1 | | | | | | | | | | | | | | | | | | | | | | | 1 | | | | | | | | | 1 | | | | | | | 7 |

| | | | | | | | | | | | | | | | | | | | | | | | | | | | | | | | | | | | | | | | | | | | | | | |
|---|---|---|---|---|---|---|---|---|---|---|---|---|---|---|---|---|---|---|---|---|---|---|---|---|---|---|---|---|---|---|---|---|---|---|---|---|---|---|---|---|---|---|---|---|---|---|---|
| | Diagnosis | | | 1 | 1 | 1 | | 1 | | 1 | 1 | 1 | | 1 | 1 | 1 | 1 | 1 | | 1 | | 1 | | 1 | 1 | | 1 | 1 | 1 | | 1 | | 1 | 1 | | 1 | | | 1 | | 1 | | 1 | 1 | 1 | | 56 |
| | Treatment | | 1 | 1 | 1 | 1 | 1 | | | 1 | 1 | 1 | 1 | 1 | 1 | 1 | 1 | 1 | 1 | 1 | 1 | | 1 | | | 1 | 1 | 1 | 1 | 1 | 1 | | 1 | 1 | 1 | | 1 | 1 | 1 | 1 | 1 | 1 | 1 | | 1 | 1 | 73 |
| | Prognosis | | | | 1 | | 1 | 1 | | | 1 | | | | | 1 | | | | | | | | 1 | | | | | | | 1 | | 1 | 1 | | | | 1 | 1 | | 1 | | 1 | | | | 22 |
| **Limitations** | | | | | | | | | | | | | | | | | | | | | | | | | | | | | | | | | | | | | | | | | | | | | | | |
| Design | | | | 1 | | 1 | 1 | 1 | 1 | 1 | | 1 | | | 1 | 1 | 1 | 1 | 1 | 1 | 1 | | | | 1 | | | 1 | | 1 | 1 | 1 | 1 | 1 | 1 | 1 | | 1 | 1 | 1 | 1 | | 1 | 1 | 1 | 1 | 67 |
| | Not optimized for the medical domain | | | 1 | | 1 | 1 | | | | | | | | 1 | 1 | 1 | | 1 | 1 | 1 | | | | 1 | | | 1 | | 1 | 1 | 1 | 1 | 1 | 1 | 1 | | 1 | 1 | 1 | 1 | | 1 | 1 | 1 | 1 | 46 |
| | | Implicit knowledge/lack of clinical context | | | | | 1 | | | | | | | | 1 | | 1 | | 1 | 1 | 1 | | | | | | | | | | | | | | | 1 | | | | | 1 | | 1 | | | | 13 |
| | | Limited clinical reasoning | | | | | | | | | | | | | | | | | | 1 | | | | | | | | | | | | | 1 | | | | | | | 1 | 1 | | 1 | 1 | | | 7 |
| | | Limited in processing/ producing medical images | | | | | | | | | | | | | | | | | | | | | | | | | | 1 | | | | | | | | | | | | | | | | | 1 | 1 | 5 |
| | | Misunderstanding of medical information/terms | | | | | | | | | | | | | | | | | | | | | | | | | | | | | | | | | 1 | | | | | | | | | | | | 7 |
| | Data | | | | | 1 | 1 | | 1 | 1 | | 1 | | | 1 | 1 | 1 | 1 | 1 | 1 | 1 | | | | 1 | | | 1 | | | 1 | 1 | 1 | 1 | | 1 | | | 1 | | 1 | | 1 | 1 | | 1 | 55 |
| | | Restricted access to internet data | | | | | | | | | | 1 | | | 1 | | | 1 | 1 | 1 | | | | | 1 | | | 1 | | | 1 | 1 | | 1 | | | | | | | | | 1 | | | | 22 |
| | | Undisclosed origin of training data | | | | 1 | 1 | | | | | 1 | | | | | | 1 | 1 | | | | | | | | | 1 | | | 1 | 1 | 1 | 1 | | | | | 1 | | | | | 1 | | 1 | 36 |
| | | Limited in reference provision/evaluation/validation | | | | | 1 | | 1 | | | 1 | | | | 1 | | | 1 | | 1 | | | | | | | 1 | | | 1 | | | 1 | | 1 | | | 1 | | 1 | | | 1 | | | 20 |
| | | Stores/processes sensitive health information | | | | | | | | | | 1 | | | | | | | 1 | | | | | | | | | | | | | 1 | | | | | | | | | | | | | | | 8 |
| | Black box | | | | | | | | 1 | | | 1 | | | | | | | 1 | | | | | | | | | 1 | | | 1 | | | | | | | | | | 1 | | | 1 | | | 12 |
| | Limited engagement/ dialogue capabilities | | | | | | | | | | | | | | | | | | | | | | | | | | | | | | | | 1 | | | | | | | | 1 | | | | | | 10 |
| | Incapable of self- | | | | | | | | 1 | 1 | | | | | | | | | | | | | | | | | | | | | | | | | | | | | | | | | | | | 1 | 4 |

| | | | | | | | | | | | | | | | | | | | | | | | | | | | | | | | | | | | | | | | | | | | | | | |
|---|---|---|---|---|---|---|---|---|---|---|---|---|---|---|---|---|---|---|---|---|---|---|---|---|---|---|---|---|---|---|---|---|---|---|---|---|---|---|---|---|---|---|---|---|---|---|---|
| | validation/correction | | | | | | | | | | | | | | | | | | | | | | | | | | | | | | | | | | | | | | | | | | | | | | |
| | Accessibility | | | | | | | | | | 1 | | | | | 1 | | 1 | | | | | | | | | 1 | | | 1 | | 1 | | | | | | | | | | 1 | 1 | | | 18 |
| | | Limited number of prompts | | | | | | | | | | | | | | | | | | | | | | | | | | | | | | | | | | | | | | | | | | 1 | | | 3 |
| | | Not freely accessible | | | | | | | | | | | | | | 1 | | | | | | | | | | | | | | | | 1 | | | | | | | | | | 1 | 1 | | | 9 |
| | | Not open source | | | | | | | | | | 1 | | | | | | 1 | | | | | | | | | 1 | | | 1 | | | | | | | | | | | | | 1 | | | 10 |
| Output | | | 1 | 1 | 1 | 1 | 1 | 1 | 1 | 1 | 1 | 1 | 1 | 1 | 1 | 1 | 1 | 1 | 1 | 1 | 1 | 1 | 1 | 1 | 1 | 1 | 1 | 1 | 1 | 1 | 1 | 1 | 1 | 1 | 1 | 1 | 1 | 1 | 1 | 1 | 1 | 1 | 1 | 1 | 1 | 1 | 86 |
| | Non-reproducible | | | | | | 1 | 1 | 1 | | | 1 | | | | 1 | | 1 | 1 | | 1 | | | | | | 1 | 1 | | | 1 | | | 1 | | 1 | 1 | | 1 | | 1 | | 1 | 1 | | | 38 |
| | | Non-deterministic | | | | | | 1 | | | | 1 | | | | | | 1 | | | | | | | | | 1 | | | | 1 | | | | | 1 | 1 | | 1 | | 1 | | 1 | | | | 24 |
| | | Non-referenceable | | | | | 1 | | 1 | | | 1 | | | | 1 | | | 1 | | 1 | | | | | | | 1 | | | 1 | | | 1 | | 1 | | | 1 | | 1 | | | 1 | | | 20 |
| | Non-comprehensive | | 1 | 1 | 1 | 1 | 1 | 1 | 1 | 1 | 1 | 1 | 1 | 1 | 1 | | 1 | 1 | 1 | 1 | 1 | 1 | 1 | 1 | 1 | 1 | 1 | 1 | 1 | 1 | 1 | 1 | 1 | 1 | 1 | 1 | | 1 | | 1 | 1 | 1 | 1 | 1 | 1 | 1 | 78 |
| | | Generic/non-personalized | | | | 1 | | | | | | | 1 | | 1 | | 1 | | | 1 | 1 | | 1 | | | | 1 | 1 | | | 1 | | 1 | | | | | 1 | | 1 | 1 | | 1 | 1 | 1 | 1 | 34 |
| | | Incomplete | 1 | 1 | 1 | | | 1 | 1 | 1 | 1 | 1 | 1 | | 1 | | 1 | 1 | 1 | 1 | 1 | 1 | 1 | | | 1 | 1 | 1 | 1 | 1 | 1 | 1 | 1 | 1 | 1 | 1 | | 1 | | 1 | 1 | 1 | 1 | 1 | 1 | 1 | 68 |
| | | Non-standard of care | | | 1 | | | 1 | 1 | | | | 1 | 1 | | | 1 | | | | 1 | | | | | | | 1 | | | 1 | | | | | 1 | | 1 | | | 1 | 1 | | | | 1 | 24 |
| | | Outdated | | | | | | | | | | | | | | | | | 1 | 1 | | | | | | | | | | | | | | | | | | | | | | | | | | | 12 |
| | | Oversimplification | | | | | | | | | | | | | 1 | | | | | 1 | 1 | | | | | | | | | | | | | | | | | | | | 1 | | | | | | 10 |
| | | Superfluous | | | | | | | 1 | | | | | | 1 | | | | | | | | 1 | | | | 1 | | | | 1 | | | | 1 | | | 1 | | | | | | | 1 | 1 | 16 |
| | | Overcautious | | | | 1 | | | | | | | | | | | | | | | | 1 | | | | | | | | | | | | | | | | | | | 1 | | | | | | 7 |
| | | Overempathic | | | | | | | | | | | | | | | | | | | | | | | | | | | | | | | | | | | | | | | | | | | | | 1 |
| | | High complexity/reading level | 1 | | | | 1 | | | | | | | 1 | 1 | | | | | | 1 | 1 | | 1 | 1 | | 1 | | | 1 | | 1 | | | | | | | | | | | | | 1 | 1 | 22 |
| | Incorrect | | | | 1 | 1 | 1 | 1 | 1 | 1 | 1 | 1 | 1 | 1 | 1 | | 1 | 1 | 1 | 1 | 1 | 1 | | 1 | 1 | 1 | 1 | 1 | 1 | 1 | 1 | 1 | 1 | 1 | 1 | 1 | 1 | 1 | 1 | 1 | 1 | 1 | 1 | 1 | 1 | 1 | 78 |
| | | Extrapolation | | | | | | | | | | | | 1 | | | 1 | | | | | 1 | | | | | | 1 | | | 1 | | | | | | | | | | | | | | 1 | 1 | 10 |
| | | Delirium | | | | 1 | | 1 | 1 | 1 | 1 | | 1 | | 1 | | 1 | 1 | 1 | 1 | 1 | 1 | | | | | 1 | 1 | 1 | | 1 | 1 | 1 | | | 1 | | | | 1 | 1 | | | | | 1 | 34 |
| | | Delusion | | | 1 | | | | 1 | 1 | | | | 1 | | | 1 | | | | | | | | | | | | | | 1 | | | | | | | | | | 1 | 1 | | | | | 14 |
| | | Hallucination | | | | | | | 1 | 1 | | 1 | 1 | | | | 1 | | 1 | | 1 | | | | | | | 1 | | | 1 | 1 | | 1 | 1 | 1 | | 1 | | | 1 | | | | 1 | 1 | 38 |
| | | Illusion | | | | | | | | | | | 1 | 1 | | | | | | | 1 | | | | 1 | | | | | | 1 | | | | | | | | | | | | | 1 | | | 12 |
| | | Confabulation | | | | 1 | | | | | | | 1 | 1 | 1 | | | | | | | | | | | | | | | | 1 | | | | | | | | | | 1 | | | | | 1 | 18 |

| | | | | | | | | | | | | | | | | | | | | | | | | | | | | | | | | | | | | | | | | | | | | | | | |
|---|---|---|---|---|---|---|---|---|---|---|---|---|---|---|---|---|---|---|---|---|---|---|---|---|---|---|---|---|---|---|---|---|---|---|---|---|---|---|---|---|---|---|---|---|---|---|---|
| | Unsafe | | | | 1 | 1 | | | 1 | 1 | | 1 | | 1 | 1 | 1 | | 1 | | 1 | 1 | | | | | | | 1 | | | 1 | 1 | 1 | | | 1 | 1 | 1 | | | | | | 1 | 1 | | 39 |
| | | Misleading | | | 1 | | | | 1 | 1 | | 1 | | 1 | 1 | 1 | | 1 | | 1 | 1 | | | | | | | | | | 1 | 1 | 1 | | | 1 | 1 | 1 | | | | | | 1 | 1 | | 34 |
| | | Harmful | | | | 1 | | | 1 | 1 | | 1 | | | 1 | | | | | 1 | 1 | | | | | | | 1 | | | 1 | 1 | 1 | | | 1 | 1 | 1 | | | | | | | | | 26 |
| | Prompt-/input dependent | | | 1 | 1 | 1 | 1 | 1 | | 1 | | 1 | | 1 | 1 | | 1 | 1 | 1 | | | | | | | | 1 | | | | 1 | | 1 | | | | | 1 | 1 | | | | | | | | 27 |
| | | Evidence | | 1 | 1 | | 1 | | | | | | | | | | 1 | 1 | | | | | | | | | | | | | | | 1 | | | | | | | | | | | | | | 7 |
| | | Complexity | | | | 1 | | | | 1 | | 1 | | | 1 | | | | 1 | | | | | | | | | | | | 1 | | | | | | | | 1 | | | | | | | | 11 |
| | | Specificity | | | | 1 | | 1 | | 1 | | 1 | | 1 | | | 1 | | | | | | | | | | | | | | | | 1 | | | | | 1 | 1 | | | | | | | | 13 |
| | | Quantity | | | | | | | | 1 | | | | | | | | | | | | | | | | | | | | | | | | | | | | | | | | | | | | | 3 |
| | | Conversation type | | | | | | | | | | | | | | | | | | | | | | | | | 1 | | | | | | | | | | | | | | | | | | | | 3 |
| | Environment-dependent | | | | | | | 1 | 1 | | | | | | | | | | | | | | | | 1 | | 1 | | | 1 | | 1 | | | 1 | | | | | | | | | | | | 16 |
| | | Target group | | | | | | 1 | | | | | | | | | | | | | | | | | | | 1 | | | 1 | | 1 | | | 1 | | | | | | | | | | | | 9 |
| | | Provider/organization | | | | | | | | | | | | | | | | | | | | | | | 1 | | | | | | | | | | | | | | | | | | | | | | 4 |
| | | Local/national medical resources | | | | | | | 1 | | | | | | | | | | | | | | | | | | | | | | | | | | | | | | | | | | | | | | 5 |
| | Bias | | | | | | | | | | | | | | | | | | | | | | | | | | | | | | | | | | | | | | | 1 | | | | | | | 6 |
| | | Language | | | | | | | | | | | | | | | | | | | | | | | | | | | | | | | | | | | | | | | | | | | | | 2 |
| | | Insurance status | | | | | | | | | | | | | | | | | | | | | | | | | | | | | | | | | | | | | | 1 | | | | | | | 1 |
| | | Underserved racial groups | | | | | | | | | | | | | | | | | | | | | | | | | | | | | | | | | | | | | | | | | | | | | 1 |
| | | Underrepresented procedures | | | | | | | | | | | | | | | | | | | | | | | | | | | | | | | | | | | | | | | | | | | | | 1 |
